# GB0139, an inhaled small molecule inhibitor of galectin-3, in COVID-19 pneumonitis: a randomised, controlled, open-label, phase 2a experimental medicine trial of safety, pharmacokinetics, and potential therapeutic value

**DOI:** 10.1101/2021.12.21.21267983

**Authors:** Erin Gaughan, Tariq Sethi, Tom Quinn, Nikhil Hirani, Andrew Mills, Annya M. Bruce, Alison MacKinnon, Vassilios Aslanis, Feng Li, Richard O’Connor, Richard A. Parker, John Norrie, James Dear, Ahsan R. Akram, Oliver Koch, Jie Wang-Jairaj, Robert J. Slack, Lise Gravelle, Bertil Lindmark, Kevin Dhaliwal

## Abstract

**Rationale:** High galectin-3 levels predict poor outcomes in patients with COVID-19. Galectin-3 activates monocytes and macrophages which are directly implicated in COVID-19 immunopathology and the cytokine storm. GB0139 is a potent thiodigalactoside galectin-3 inhibitor and may reduce the severe effects of the disease. We report safety and pharmacokinetics and pharmacodynamics of the inhaled galectin-3 inhibitor, GB0139, and assess clinical outcomes and key systemic inflammatory biomarkers in hospitalised patients with COVID-19 (ClinicalTrials.gov/EudraCT identifier: NCT04473053/2020-002230-32).

**Methods:** Adults with COVID-19 requiring oxygen, and with pneumonitis on x-ray, were randomised to receive standard of care (SOC; including dexamethasone; n=21) or SOC plus 10 mg GB0139 twice daily for 48 hours, then once daily for ≤14 days (n=20).

**Results:** Patients aged 27–87 years were enrolled from July 2020; the final patient completed the 90-day follow-up in April 2021. GB0139+SOC was well tolerated with no treatment-related serious adverse events reported. Incidences of adverse events were similar between treatment arms (40 with GB0139+SOC *vs* 35 with SOC). Plasma GB0139 was measurable in all patients after inhaled exposure, with moderate interpatient variability, and demonstrated target engagement with decreased circulating galectin (overall treatment effect post-hoc over days 2–7: p=0·0099 *vs* SOC). Rate of decline in fraction of inspired oxygen (%) requirement was significantly greater in the GB0139+SOC arm with a posterior mean difference of -1·51 (95% highest posterior density: -2·90, -0·189) versus SOC. Plasma levels of biomarkers associated with inflammation, coagulopathy, major organ function and fibrosis showed a downward trend versus SOC.

**Conclusions:** GB0139+SOC was well tolerated and achieved clinically relevant plasma concentrations and target engagement. This, and the reduction in markers associated with inflammatory, coagulation, fibrosis, and reduction in inspired oxygen (%) over SOC alone, indicates the therapeutic potential for inhaled GB0139 in hospitalised patients with COVID-19.

## Introduction

Severe acute respiratory syndrome coronavirus 2 (SARS-CoV-2) and the disease it causes (coronavirus disease 2019 [COVID-19]) has put major stress on the world’s healthcare services, and despite vaccination, is likely to remain a problem for the foreseeable future.^1^ Severe morbidity and mortality from COVID-19 is predominantly a consequence of immunopathology in response to virus infection,^2^ characterised by aberrant monocyte and macrophage-driven acute respiratory distress syndrome (ARDS), T cell depletion, inflammation-driven thrombosis, and fibrotic lung damage.^3-6^

Dexamethasone has been shown to reduce mortality in hospitalised patients who require supplemental oxygen with the greatest effect observed in those requiring mechanical ventilation.^7^ An incremental benefit has also been demonstrated with anti-interleukin (IL)-6 monoclonal antibodies (eg, tocilizumab, a humanised monoclonal antibody),^8,9^ and the broad-spectrum antiviral, remdesivir, may improve time to recovery.^10^ Despite the approval of these agents as standard of care (SOC) for COVID-19, mortality is still high due to ARDS hypoxia, microvascular thrombosis and multiorgan failure.^10,11^ It is clear that there is still an urgent need to identify further effective and tolerable therapies.

Galectin-3 is a mammalian, ß-galactoside-binding lectin, which is an important regulator of immune homeostasis and is highly upregulated following lung injury.^12-15^ Galectin-3 has been shown to drive a pro-inflammatory cytokine storm following co-infection of pneumococcus and H1N1 avian influenza virus,^16^ with upregulation of macrophage IL-1β production.^17^ In alveolar epithelial cells, galectin-3 increases tumour necrosis factor (TNF), IL-6, and IL-8 release during *Streptococcus pneumoniae* and H1N1 influenza co-infection, and enhances the pathogenic effects by promoting excessive host inflammatory responses via macrophage NLRP3 inflammasome activation.^16,17^ Galectin-3 also acts as a ligand for Toll-like receptor 4,^18,19^ promoting monocyte and macrophage recruitment to damaged tissue.^20^ Recently, galectin-3 has been identified as a potential marker of lung damage, an independent predictor of poor outcomes in patients with COVID-19,^21^ and a possible therapeutic target.^6,22^

Highly elevated levels of galectin-3, IL-1, IL-6, and TNF-α have been observed in patients with severe COVID-19, compared with those who have moderate disease.^21,23^

As well as direct galectin-3 inhibition, anti-galectin-3 therapy can block macrophage and dendritic cell secretion of cytokines IL-1, IL-6, and TNF-α, and may attenuate the cytokine storm and other inflammatory COVID-19-associated consequences.^21,22,24^ Furthermore, galectin-3 inhibition may block venous and inflammation-driven thrombosis.^25^

Galectin-3 has been identified as a therapeutic target in patients with idiopathic pulmonary fibrosis (IPF) supported by both in vitro and in vivo pre-clinical models.^14^ In a phase 1/2a trial, inhaled GB0139, a potent thiodigalactoside galectin-3 inhibitor, suppressed galectin-3 expression and decreased key plasma biomarkers associated with IPF disease progression.^26^ These initial findings support the hypothesis that inhaled GB0139 may counteract the SARS-CoV-2-associated galectin-3 disease drive and reduce the severe effects of COVID-19, including the development of post-viral pulmonary fibrosis. We therefore sought to investigate the safety and pharmacokinetics (PK) and pharmacodynamics (PD) of inhaled GB0139 in hospitalised patients with COVID-19 pneumonitis, prior to evaluating clinical efficacy in larger trials. Biological markers of efficacy were also explored.

## Methods

### Study design

DEFINE (University of Edinburgh COVID-19 DEFINE trial, 2020; ClinicalTrials.gov identifier: NCT04473053, EurdraCT number: 2020-002230-32) was a randomised, controlled, open-label, parallel-group, experimental medicine platform study evaluating safety and PK and PD of GB0139 in patients with COVID-19; it was not powered for clinical efficacy. A study cohort size of approximately 20 patients per group was chosen.

After initial screening for eligibility, patients were randomly assigned (1:1:1) to receive GB0139 plus SOC, nafamostat plus SOC, or SOC alone. Randomisation was carried out by a member of the research team using a centralised web-based service provided by the Edinburgh Clinical Trials Unit (Usher Institute, University of Edinburgh, Edinburgh, UK). A minimisation procedure based on sex, age, body mass index, and a history of diabetes was used for randomisation and incorporated a 20% random element, meaning that the treatment arm selected by the minimisation procedure was switched with a probability of 20%. Patients, investigators, and treating clinicians were unaware of treatment allocations before they were assigned to each individual patient. SOC included dexamethasone, tocilizumab, antibiotics, low molecular weight heparin, and remdesivir, and was determined by the treating physician. For this analysis, we report data from the GB0139+SOC and SOC cohorts only. Results for the nafamostat arm of the study have been published.^27^

GB0139 was administered as an inhaled formulation of 5 mg x 2 (10 mg total) twice daily (BID) via a dry powder inhaler (Plastiape™, Berry Bramlage, Lohne, Germany) for the first 48 hours, followed by 10 mg once daily (QD) for up to 14 days in total, or until discharge from hospital or withdrawal from the trial. There were no changes to the dosing regime and clinical assessments prior to trial commencement.

### Ethics

The study was conducted in accordance with the Declaration of Helsinki and the principles of the International Conference on Harmonization Tripartite Guideline for Good Clinical Practice. Independent ethics committee approval (Scotland A Research Ethics Committee: 20/SS/0066) was obtained prior to initiation of the study. The independent Data Monitoring Committee scrutinised accumulating data.

### Study participants

Eligible patients were aged ≥16 years, had polymerase chain reaction-confirmed COVID-19 infection, and were hospitalised with breathlessness requiring oxygen and evidence of pneumonitis on x-ray. Full inclusion and exclusion criteria are provided in table 1.

**Table 1:** Inclusion and exclusion criteria for patient eligibility for study participation.

| Inclusion criteria | Exclusion criteria |
| --- | --- |
| - COVID-19 positive test result within the last 14 days<br>- Hospitalised with breathlessness requiring oxygen and evidence of pneumonitis on x-ray<br>- Provision of informed consent from the patient or representative<br>- Aged at least 16 years<br>- If the patient is of child-bearing potential, the patient, and their partner(s), agree to use medically accepted double-barrier methods of contraception during the study and for at least 90 days after termination of study therapy | - Current or recent history, as determined by the Investigator, of severe, progressive, and/or uncontrolled cardiac disease (NYHA class IV), uncontrolled renal disease (eGFR <30 mL/min/1.73 m<sup>2</sup>), severe liver dysfunction (ALT >5x ULN) or anaemia (Hb <80 g/L)<br>- Women who are pregnant or breastfeeding<br>- Participation in another clinical trial of an investigational medicinal product<br>- Known hypersensitivity to the IMP or excipients (eg, lactose)<br>- Concomitant use of treatments for COVID-19 that are not recognised as locally approved standard care<br>- Significant electrolyte disturbance (hyperkalaemia K<sup>+</sup> >5.0 mmol/L or hyponatraemia Na<sup>+</sup> <120 mmol/L)<br>- Currently receiving potassium sparing diuretics that cannot be reasonably withheld<br>- Currently receiving anticoagulation or antiplatelet agents that cannot be reasonably withheld if randomised to receive nafamostat<br>- Patients (or their partners) planning on donating sperm/eggs during the trial period<br>- Ongoing dialysis<br>- History of serious liver disease (Child Pugh score >10)<br>- Severe uncontrolled diabetes mellitus<br>- In the Investigator's opinion, patient is unwilling or unable to comply with drug administration plan, laboratory tests or other study procedures |

ALT=alanine aminotransferase. AST=aspartate transaminase. COVID-19=coronavirus disease 2019. eGFR=estimate glomerular filtration rate. Hb=haemoglobin.
IMP=investigational medicinal product. NYHA=New York Heart Association. ULN=upper limit of normal.

### Objectives

The primary objective was to evaluate the safety of inhaled GB0139 as add-on therapy to SOC in patients with COVID-19. Secondary objectives included assessments of PK and PD properties of GB0139. Key exploratory biomarkers were assessed during treatment, as was potential for clinical efficacy.

#### Safety and tolerability outcomes

Safety was assessed daily using vital signs (supine blood pressure, pulse, oxygen saturation, tympanic temperature, and respiratory rate), 12-lead electrocardiogram (ECG), physical examination, and laboratory safety screen (haematology, coagulation, biochemistry). All adverse events (AE) and concomitant medications were recorded from the time of the screening visit until discharge, withdrawal, or death.

#### Pharmacokinetics

Blood samples for PK assessments were drawn pre-dose on days 1 (first day of dosing), 3, 5, 8, and 11 (+/- 1 day) for the determination of trough plasma GB0139 concentrations. Blood plasma samples were analysed by Simbec Orion (Merthyr Tydvill UK) using a validated high performance liquid chromatography method with tandem mass spectrometry detection. The lower limit of quantitation (LLOQ) was 0·5 ng/mL.

#### Flow cytometry

Inflammatory cell phenotypes were analysed by fluorescence-activated cell sorting (FACS). Methods are described in the Supplementary Appendix, and gating strategies and list of antibodies used are available in figure S1 and table S1, respectively.

#### Biomarkers

Biomarker assessments were performed daily, subject to patient tolerance. Pre-selected relevant biomarkers of COVID-19 (IL-1β, IL-6, IL-8, TNF-α, C–C motif chemokine ligand 2, IL-17A, IL-10, C-X-C motif chemokine ligand 10 [CXCL10], granulocyte-macrophage colony-stimulating factor (CSF), amphiregulin and IL-1rα)^28^ were analysed using the ELLA platform. Relevant inflammatory biomarkers were analysed (C-reactive protein [CRP], neutrophil/lymphocyte ratio [NLR]) by routine clinical measurements, and lymphocyte and monocyte phenotype determined by flow cytometry. Also investigated were coagulopathy-related biomarkers (D-dimer levels, platelets, fibrinogen/platelet ratio [FPR] and activated partial thromboplastin time [aPTT]),^29^ and markers of organ function/acute phase response (fraction of inspired oxygen [FiO_2_], creatine kinase, aspartate aminotransferase [AST], gamma-glutamyltransferase [GGT] and alkaline phosphatase).^30^ Plasma biomarkers known to be involved in pulmonary fibrosis^26^ were analysed by ELISA (chitinase-3-like protein 1 [YKL-40] and plasminogen activator inhibitor-1 [PAI-1]) (R&D Systems, MN, USA).

### Statistical analysis

The analysis population included all patients, with the exception of those randomised to receive GB0139+SOC but who did not receive any study drug. Analysis was undertaken without any formal imputation of missing data.

Bayesian Generalised Linear Mixed Effects models (GLMM) were fitted to continuous safety outcomes, with baseline and trial arm as explanatory variables, and a random effect for patient. Day of measurement post-randomisation was also included as a categorical factor variable. Non-informative flat priors were used. Results were reported as posterior mean differences and highest posterior density (HPD) intervals.

To evaluate the change from baseline values for key secondary variables (FiO_2_, National Early Warning score 2 [NEWS2],^31^ World Health Organisation score), a Bayesian GLMM was fitted including the following explanatory variables: baseline, patient (as a random effect), day of measurement post-randomisation (as a continuous variable), trial arm (GB0139+SOC/SOC), and an interaction term for day of measurement and trial arm. Therefore, we assessed whether changes in secondary variables over time significantly differed by treatment arm by means of the interaction term. Pearson correlation was used to evaluate whether there was statistical evidence for a linear relationship among galectin-3 levels and biomarkers of interest. The p-value for target engagement was calculated using an Analysis of Covariance model adjusting for treatment and baseline galectin-3. This was done using information from days 2–7 and with assessment day as a repeated measure. The p value assessed the significance of the variable treatment in the model.

For AEs, a Bayesian logistic regression model was used to compare the odds of “at least one” AE between trial arms; and a Bayesian Poisson model was fitted to the total number of AEs per patient to compare rates of AEs. Trial arm was the only explanatory variable in the models, with no other covariates. All statistical models were fitted using SAS software (SAS Institute Inc., Cary, NC, USA).

## Results

### Patient disposition and baseline characteristics

Patients were recruited from September 2020 to February 2021. The study population comprised 43 patients aged 27–87 years who were admitted to the Royal Infirmary of Edinburgh and Western General Hospital, Edinburgh, with symptomatic COVID-19; 22 patients were randomised to receive GB0139+SOC and 21 to receive SOC alone (figure 1). Two patients assigned to the GB0139+SOC arm did not receive any study drug and were excluded from the analyses. One patient missed the loading dose. Full baseline demographics and disease characteristics are provided in table 2. No patient in the study population was a care home resident.

**Table 2:**
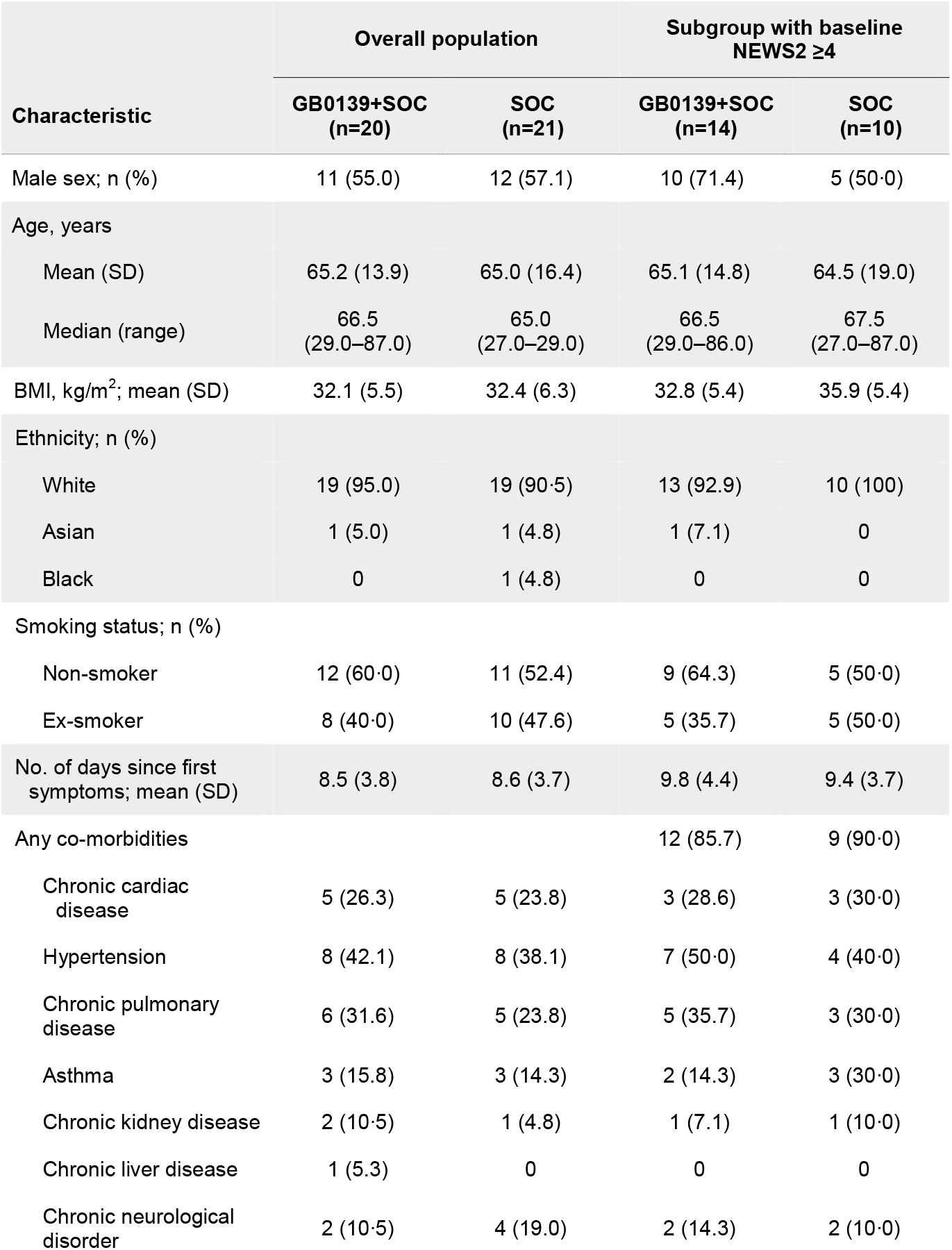
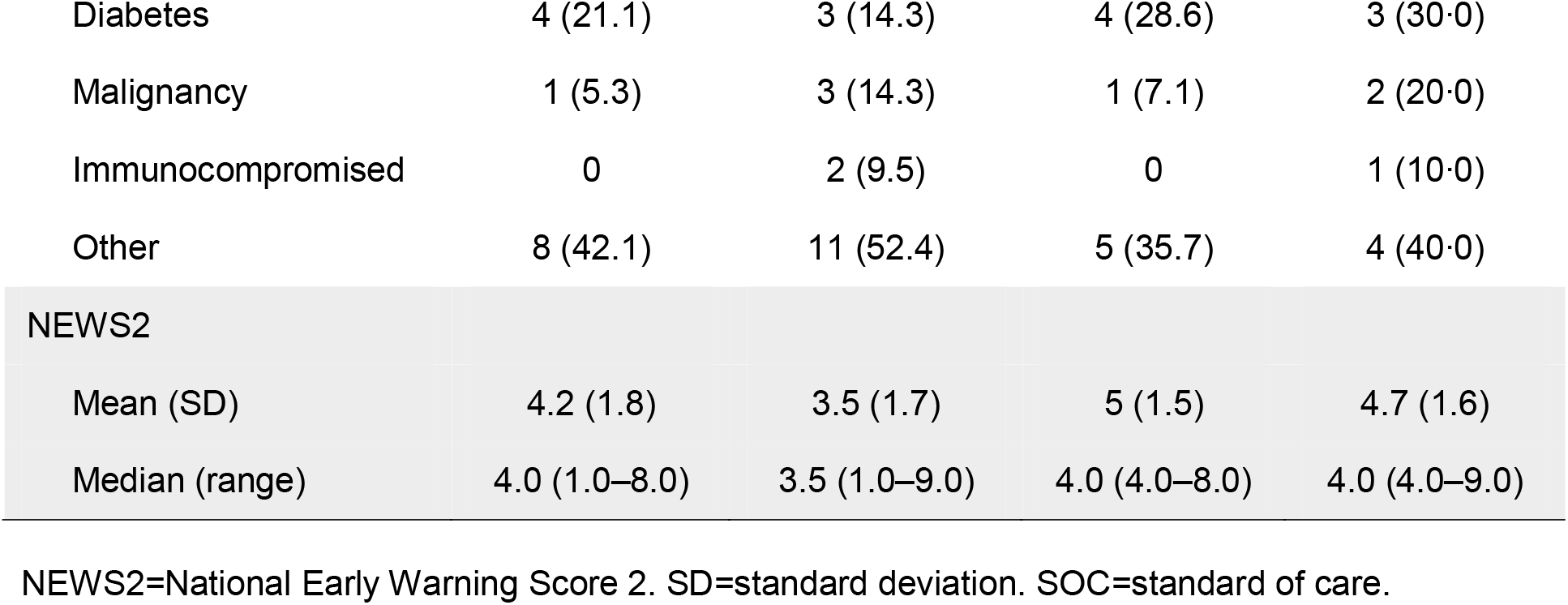
Patient demographics and disease characteristics at study entry.

**Figure 1:**
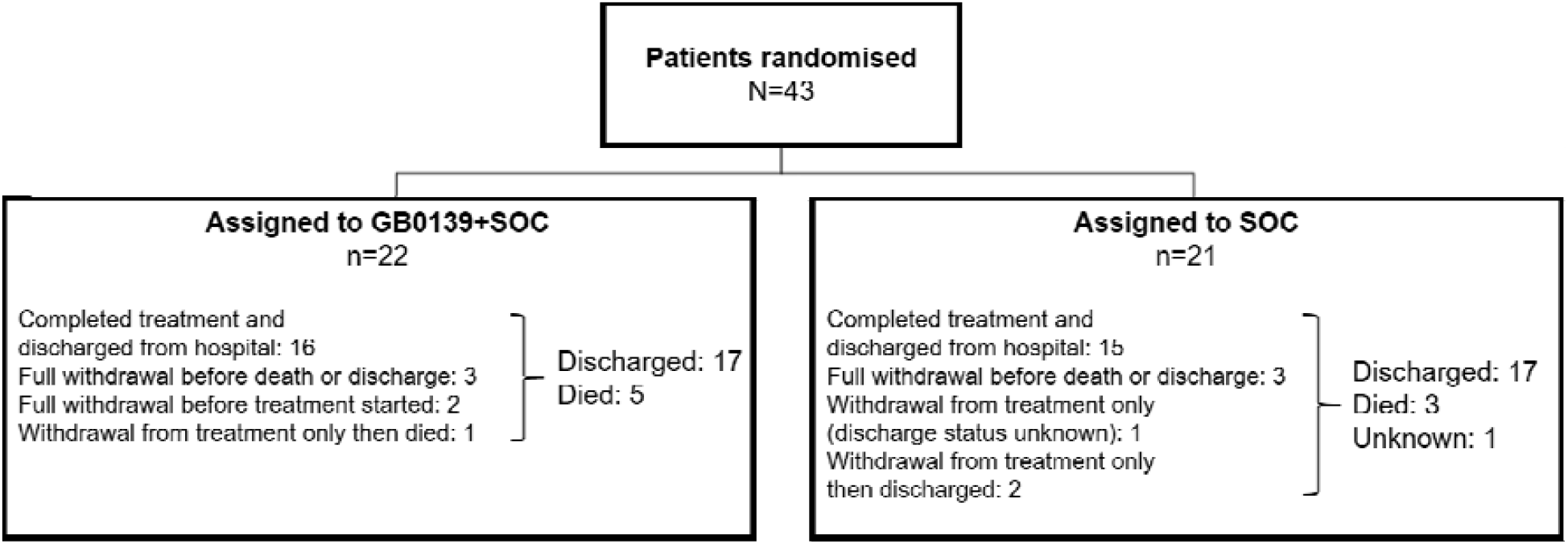
Patient disposition. SOC=standard of care.

### Safety and tolerability of GB0139

There were no safety or tolerability concerns associated with inhaled administration of GB0139, and no statistically significant differences observed in haematological and biochemical safety laboratory outcomes or vital signs (table S2). The total number of AEs in patients who received GB0139+SOC was 40 compared with 35 in those who received SOC. The total number of patients who experienced at least one AE was 14 (70·0%) in the GB0139+SOC group versus 12 (57·1%) in the SOC group (table 3). The odds ratio (95% confidence interval [CI]) of ≥1 AE occurring between trial arms was 1·83 (0·48–6·81) and the rate ratio (95% CI) between arms for the number of AEs per patient was 1·20 (0·75–1·89). Although there were three serious AEs recorded from one patient following GB0139+SOC treatment and hospital discharge (small bowel obstruction and paracetamol overdose both requiring hospital admission, and worsening of hyponatraemia), these were considered unrelated to study treatment and occurred in the same patient post-hospital discharge (for COVID-19).

**Table 3:** Adverse events in the safety population.

| Study group | Total population | | Subgroup with baseline NEWS2 $\geq 4$ | |
| --- | --- | --- | --- | --- |
|  | GB0139+SOC (n=20) | SOC (n=21) | GB0139+SOC (n=14) | SOC (n=10) |
| Any AE, n (%) | 14 (70.0) | 12 (57.1) | 10 (71.4) | 4 (40.0) |
| Any serious AE, n (%) | 1 (5.0) | 0 (0.0) | 0 (0.0) | 0 (0.0) |

AE=adverse event. NEWS2=national early warning score 2. SOC=standard of care. \*Three serious adverse events were reported for one patient in the GB0139 study group; small bowel obstruction, paracetamol overdose and worsening of hyponatraemia.

Amongst patients who received GB0139+SOC, five reported an AE considered by the clinical study team to be possibly related to treatment. These comprised nausea (due to the inhaler powder), prolonged QTc on ECG, sore throat, oral thrush, and hair loss (all mild severity). One patient assigned to SOC reported mild atrial fibrillation that was possibly related to treatment.

### Plasma concentrations of GB0139

After an initial peak of 19·94 ng/mL on day 3, consistent with the higher 10 mg BID dosing for the first 2 days, geometric mean trough plasma concentrations of GB0139 decreased to 7·81 ng/mL on day 5 and had reached steady-state by day 8 (6·01 ng/mL). Similar trough concentrations were measured at the end of the PK assessment period on day 11 (5·50 ng/mL). The apparent decrease of plasma concentrations between days 3 and 5 was due to the two-fold lower daily dose administered from day 3 onwards (10 mg QD from day 3 *vs* 10 mg BID for the first 48 hours). Ranges of individual values overlapped between days 5, 8, and 11, and interindividual variability on trough plasma concentrations was moderate, with coefficients of variation ranging from 39% to 57%, indicating consistent exposure between patients and across days (table 4). A previous study in patients with IPF found GB0139 treatment resulted in reduced levels of biomarkers associated with fibrosis.^14,26^ To pre-empt GB0139 activity in patients with COVID-19 using their plasma levels, 10 mg GB0139 trough concentrations were plotted against values previously derived from the aforementioned patients with IPF. Inhaled GB0139 achieved plasma concentrations in patients with COVID-19 comparable to that previously observed in patients with IPF (unpublished data on file; table S3) with individual ranges overlapping between the two populations (figure S2).

**Table 4:** Trough (pre-dose) plasma GB0139 concentrations (ng/mL) in patients with COVID-19.

|  | Baseline | Day 3 | Day 5 | Day 8 | Day 11 |
| --- | --- | --- | --- | --- | --- |
| n | 18 | 20 | 12 | 9 | 4 |
| Arithmetic mean (SD) | BLQ | 22.91 (12.03) | 8.58 (4.60) | 6.66 (3.80) | 5.85 (2.30) |
| CV, % | – | 53 | 54 | 57 | 39 |
| Geometric mean | – | 19.94 | 7.81 | 6.01 | 5.50 |
| Median (min–max) | BLQ (BLQ–BLQ) | 18.28 (7.45–51.21) | 7.32 (3.94–21.62) | 5.27 (3.87–15.99) | 5.70 (3.22–8.81) |

BLQ=below the limit of quantitation (0.5 ng/mL). CV=coefficient of variation. SD=standard deviation.

### GB0139 effect on biomarkers

Analyses of GB0139 on biomarkers were all post-hoc with 20 patients included in each treatment arm. There was a decrease in the mean serum concentration of galectin-3 over time with GB0139+SOC treatment (overall treatment effect post-hoc *vs* SOC over days 2–7: p=0·0099), indicating target engagement activity. Of note, overall treatment effect, using all available data to day 16 was also significant for GB0139+SOC versus SOC (post-hoc p=0·0001) however the last day for data availability in the SOC arm was day 14 (figure 2).

**Figure 2:**
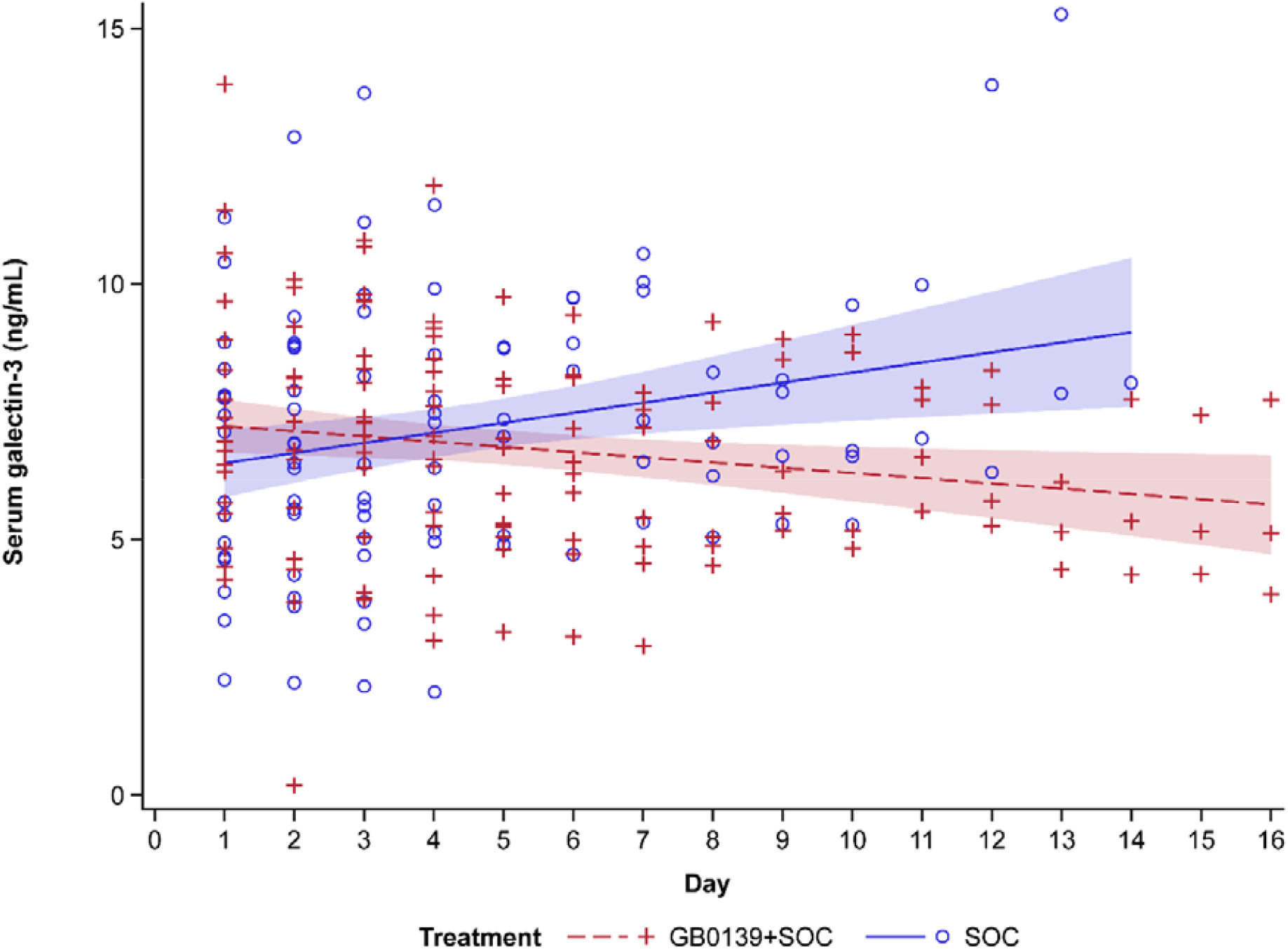
Individual patient serum galectin-3 concentrations from all patients receiving GB0139+SOC (red) or SOC (blue) Linear regression analysis showing line of best fit with 95% CI (shaded area). CI=confidence interval. SOC=standard of care.

#### Biomarkers of inflammation

Although GB0139+SOC-treated patients had higher baseline CRP levels than those receiving SOC (115 *vs* 45 mg/L, respectively), a greater downward trend in mean change from baseline CRP was observed in the former group (figure 3A). Similarly, patients receiving GB0139+SOC showed a trend in decreasing lactate dehydrogenase and NLR compared with patients receiving SOC (figure 3B and C). This suggests GB0139+SOC had a greater effect on reducing inflammation than SOC alone.

**Figure 3:**
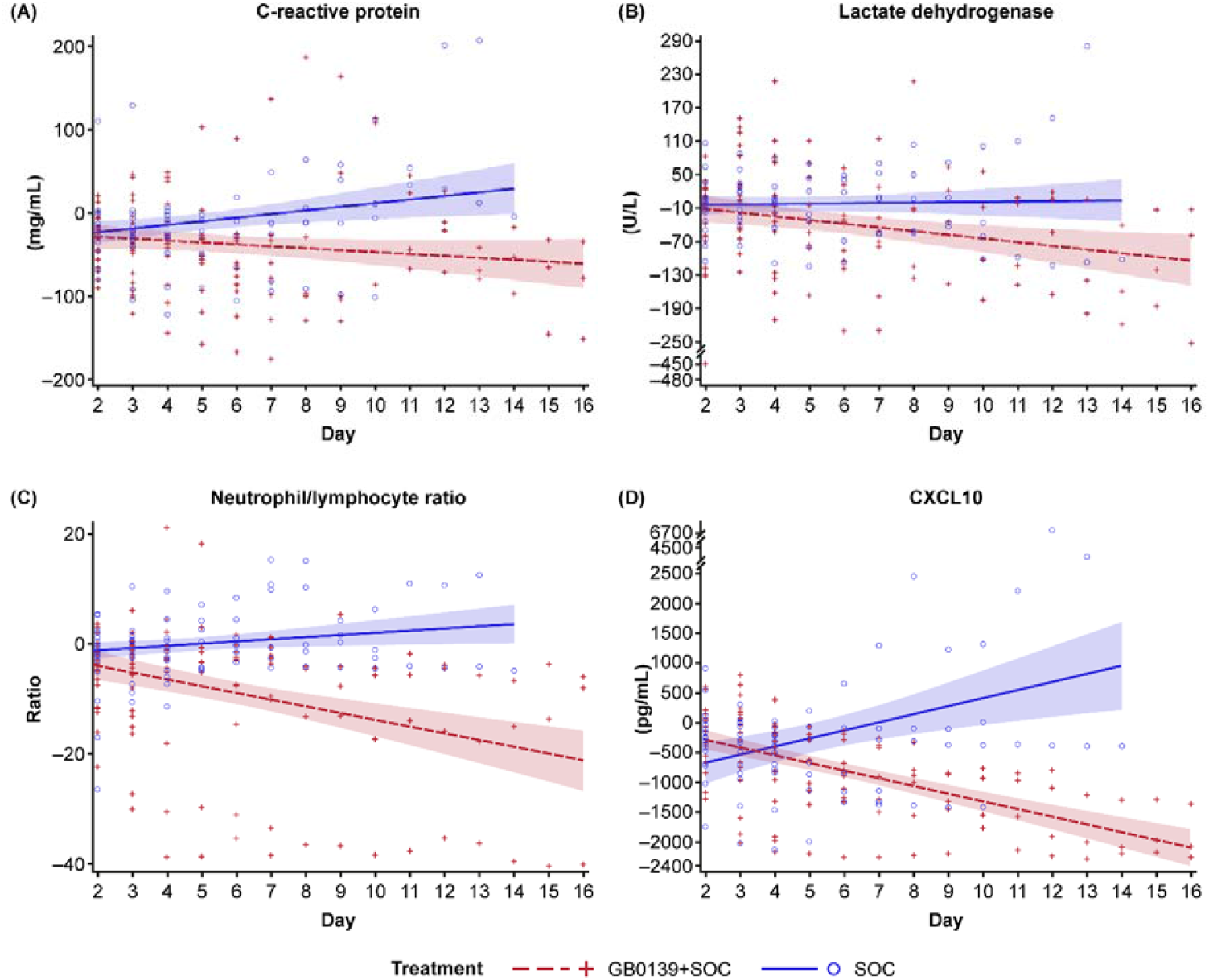
Absolute change from baseline in markers of inflammation in the overall population: (A) C-reactive protein, (B) lactate dehydrogenase, (C) neutrophil/lymphocyte ratio, and (D) CXCL10. Individual patient-level data from all those who received GB0139+SOC (red) or SOC (blue): linear regression analysis showing line of best fit with 95% CI (shaded area). CI=confidence interval. CXCL10=C-X-C motif chemokine ligand 10. SOC=standard of care.

Patients receiving GB0139+SOC also showed a downward trend in CXCL10 levels compared with those receiving SOC (figure 3D). These reductions in CXCL10 levels had a moderate, positive correlation with reductions in galectin-3 levels in the GB0139+SOC treatment group (p<0·05 at days 2, 3 and 4, with Pearson correlation coefficients of 0·47, 0·65, and 0·55, respectively; table 5). There also appeared to be downward trends in the mean change from baseline of cytokines IL-10, IL-6, and TNF (up to day 7) in the GB0139+SOC group (figure S3).

**Table 5:**
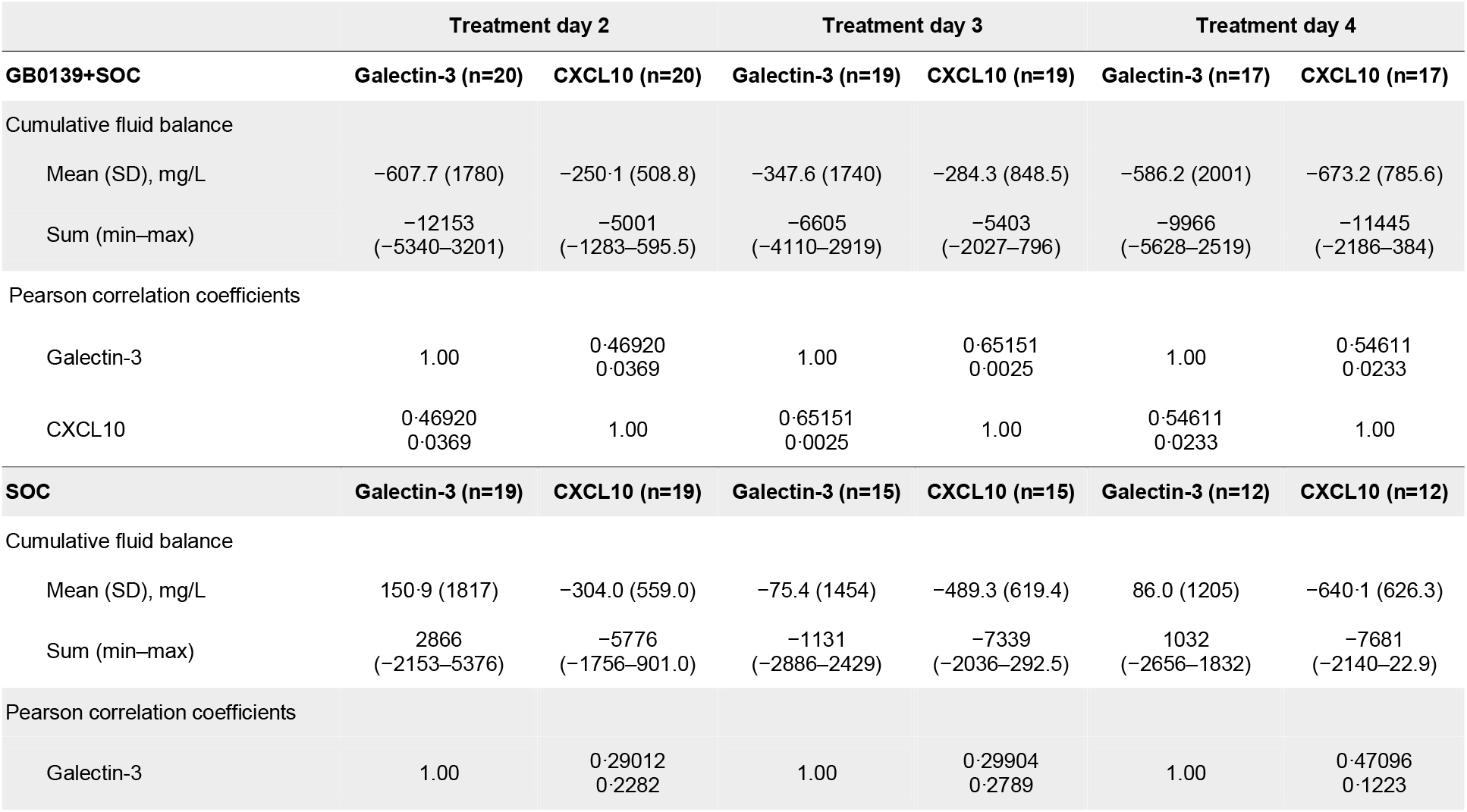
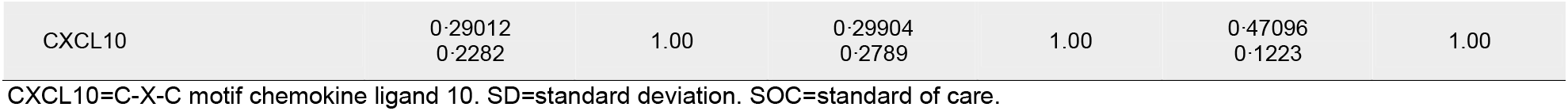
**Trend in CXCL10 reduction and a correlation between change from baseline in galectin-3 and CXCL10 upon treatment with GB0139+SOC or SOC alone**

|  | Treatment day 2 |  | Treatment day 3 |  | Treatment day 4 |  |
| --- | --- | --- | --- | --- | --- | --- |
| **GB0139+SOC** | **Galectin-3 (n=20)** | **CXCL10 (n=20)** | **Galectin-3 (n=19)** | **CXCL10 (n=19)** | **Galectin-3 (n=17)** | **CXCL10 (n=17)** |
| Cumulative fluid balance |  |  |  |  |  |  |
| Mean (SD), mg/L | −607.7 (1780) | −250.1 (508.8) | −347.6 (1740) | −284.3 (848.5) | −586.2 (2001) | −673.2 (785.6) |
| Sum (min–max) | −12153<br>(−5340–3201) | −5001<br>(−1283–595.5) | −6605<br>(−4110–2919) | −5403<br>(−2027–796) | −9966<br>(−5628–2519) | −11445<br>(−2186–384) |
| Pearson correlation coefficients |  |  |  |  |  |  |
| Galectin-3 | 1.00 | 0.46920<br>0.0369 | 1.00 | 0.65151<br>0.0025 | 1.00 | 0.54611<br>0.0233 |
| CXCL10 | 0.46920<br>0.0369 | 1.00 | 0.65151<br>0.0025 | 1.00 | 0.54611<br>0.0233 | 1.00 |
| **SOC** | **Galectin-3 (n=19)** | **CXCL10 (n=19)** | **Galectin-3 (n=15)** | **CXCL10 (n=15)** | **Galectin-3 (n=12)** | **CXCL10 (n=12)** |
| Cumulative fluid balance |  |  |  |  |  |  |
| Mean (SD), mg/L | 150.9 (1817) | −304.0 (559.0) | −75.4 (1454) | −489.3 (619.4) | 86.0 (1205) | −640.1 (626.3) |
| Sum (min–max) | 2866<br>(−2153–5376) | −5776<br>(−1756–901.0) | −1131<br>(−2886–2429) | −7339<br>(−2036–292.5) | 1032<br>(−2656–1832) | −7681<br>(−2140–22.9) |
| Pearson correlation coefficients |  |  |  |  |  |  |
| Galectin-3 | 1.00 | 0.29012<br>0.2282 | 1.00 | 0.29904<br>0.2789 | 1.00 | 0.47096<br>0.1223 |
| CXCL10 | 0.29012<br>0.2282 | 1.00 | 0.29904<br>0.2789 | 1.00 | 0.47096<br>0.1223 | 1.00 |

CXCL10=C-X-C motif chemokine ligand 10. SD=standard deviation. SOC=standard of care.

In the current study, 40% of patients had mild disease (defined as NEWS2 ≤3) but there was a marked difference in the clinical severity between GB0139+SOC and SOC groups, with 30% of patients randomised to GB0139+SOC identified as having low clinical risk at baseline (NEWS2 ≤3) compared with 50% in the SOC group. Similarly, 20% of patients in the GB0139+SOC arm were of high clinical risk at baseline (NEWS2 ≥6), versus 5% in the SOC arm. As patients with severe disease may benefit more from lowered galectin-3, a post-hoc subgroup analysis of patients with a baseline NEWS2 ≥4 was undertaken. The downward trends in cytokines CXCL10, IL-10, IL-6, and TNF were particularly evident in this subgroup (figure 4A–D), and analysis of lymphocytes by flow cytometry showed that while total lymphocyte numbers remained steady in the SOC arm, there was a tendency for these to increase with GB0139+SOC. Markers of CD4 T-cell activation were similar across treatment groups. Activation markers on CD8 T cells appeared stable to day 4 in both groups, but at day 7 were higher with SOC versus GB0139+SOC. There was a consistent decrease in exhaustion markers for both CD4 and CD8 in patients treated with GB0139+SOC compared with an increase in exhaustion markers seen in those receiving SOC. There was a trend towards a higher percentage of B cells with GB0139+SOC treatment compared with SOC (figure 5).

**Figure 4:**
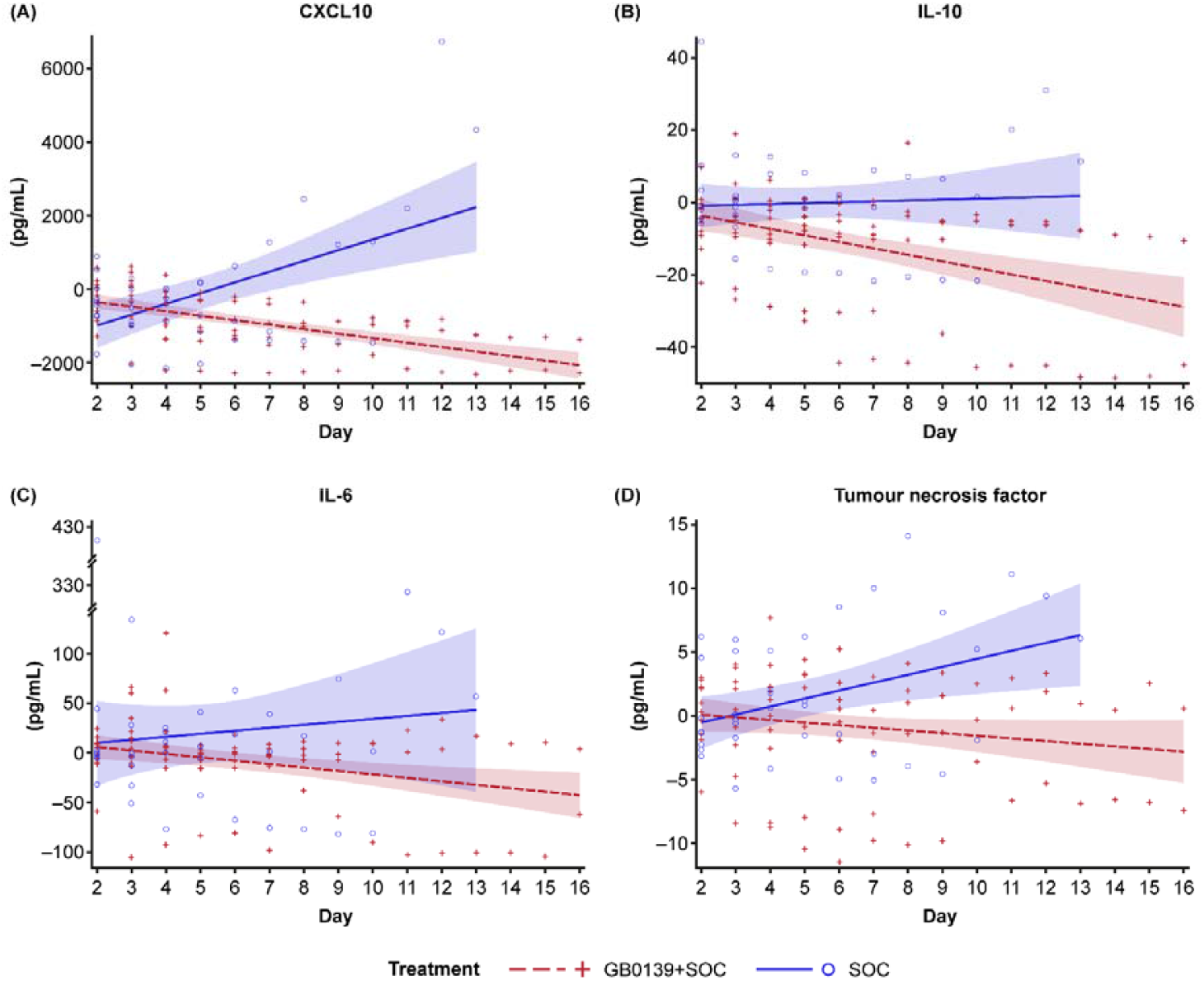
Change from baseline in markers of inflammation in the population with baseline NEWS2 ≥4: patient level data for (A) CXCL10, (B) IL-10, (C) IL-6, and (D) tumour necrosis factor. Individual patient-level data from all those who received GB0139+SOC (red) or SOC (blue): linear regression analysis showing line of best fit with 95% CI (shaded area). CI=confidence interval. CXCL10=C-X-C motif chemokine ligand 10. IL=interleukin. NEWS2=national early warning score 2. SOC=standard of care.

**Figure 5:**
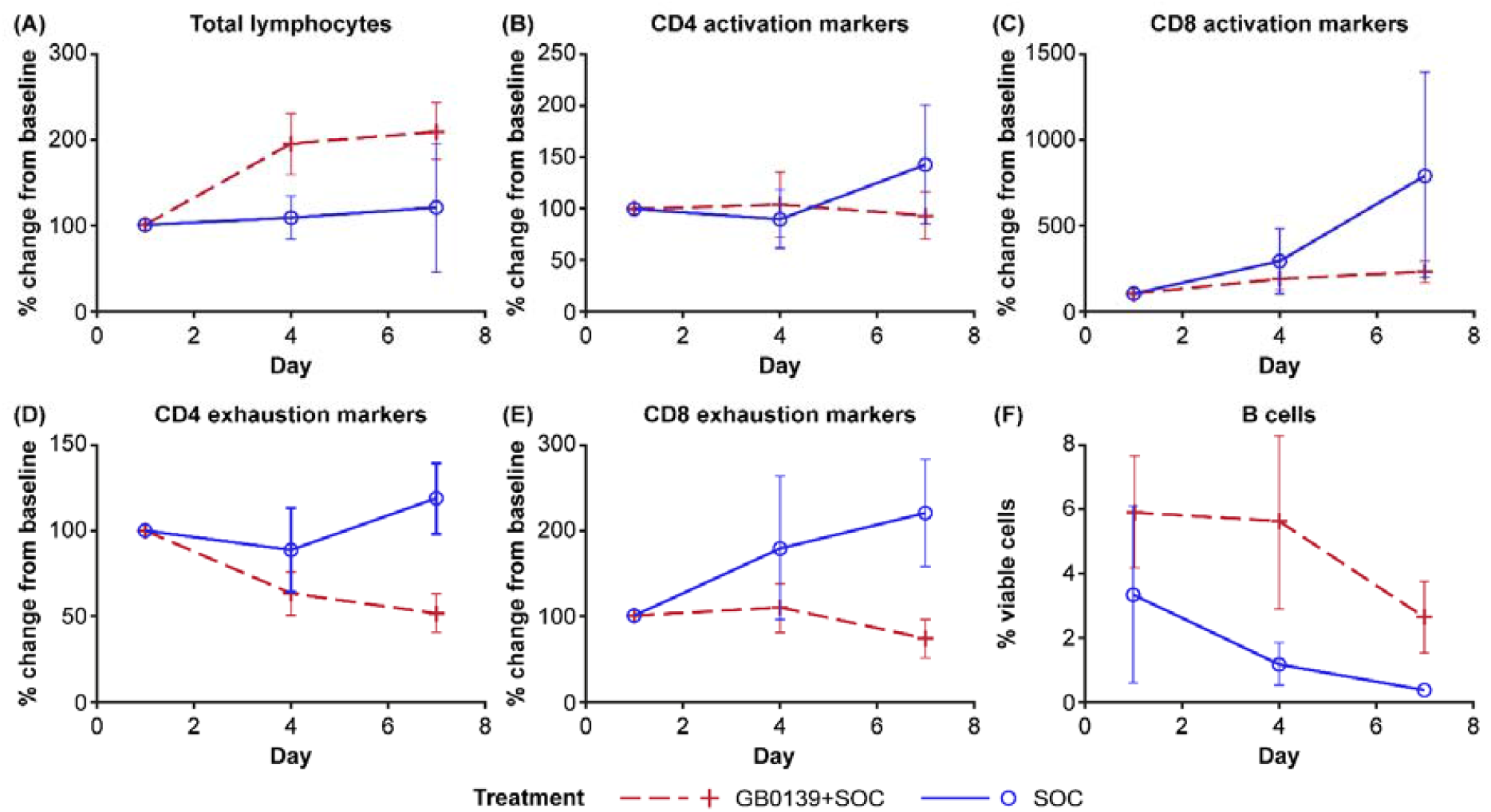
Effect of GB0139+SOC (red) and SOC (blue) on circulating lymphocytes in the population with baseline NEWS2 ≥4: (A) total lymphocytes, (B) CD4 T cell activation markers, (C) CD8 T cell activation markers, (D) CD4 T cell exhaustion markers, (E) CD8 T cell exhaustion markers, and (F) B cells. Data are expressed as mean (±SEM) % change from baseline or % viable cells. NEWS2=national early warning score 2. SEM=standard error of the mean. SOC=standard of care.

All biomarkers were assayed as planned. Cytokines IL-1b and granulocyte-macrophage CSF are not reported due to low or undetectable levels (below the LLOQ). IL-1rα, IL-8, IL-17, and amphiregulin were measured but no differences were noted between treatment arms.

#### Biomarkers of coagulopathy

Patients receiving GB0139+SOC had consistently low levels of D-dimer compared with patients receiving SOC alone (figure 6A). Lower levels of FPR and a downward trend over time were observed in FPR with GB0139+SOC while an upward trend was apparent with SOC (figure 6B). Furthermore, an upward trend in platelet count and aPTT was seen in GB0139+SOC-treated patients and a downward trend in patients receiving SOC (figure 6C and D). These results suggest that patients receiving GB0139+SOC may have a lower likelihood of thrombosis versus SOC alone.

**Figure 6:**
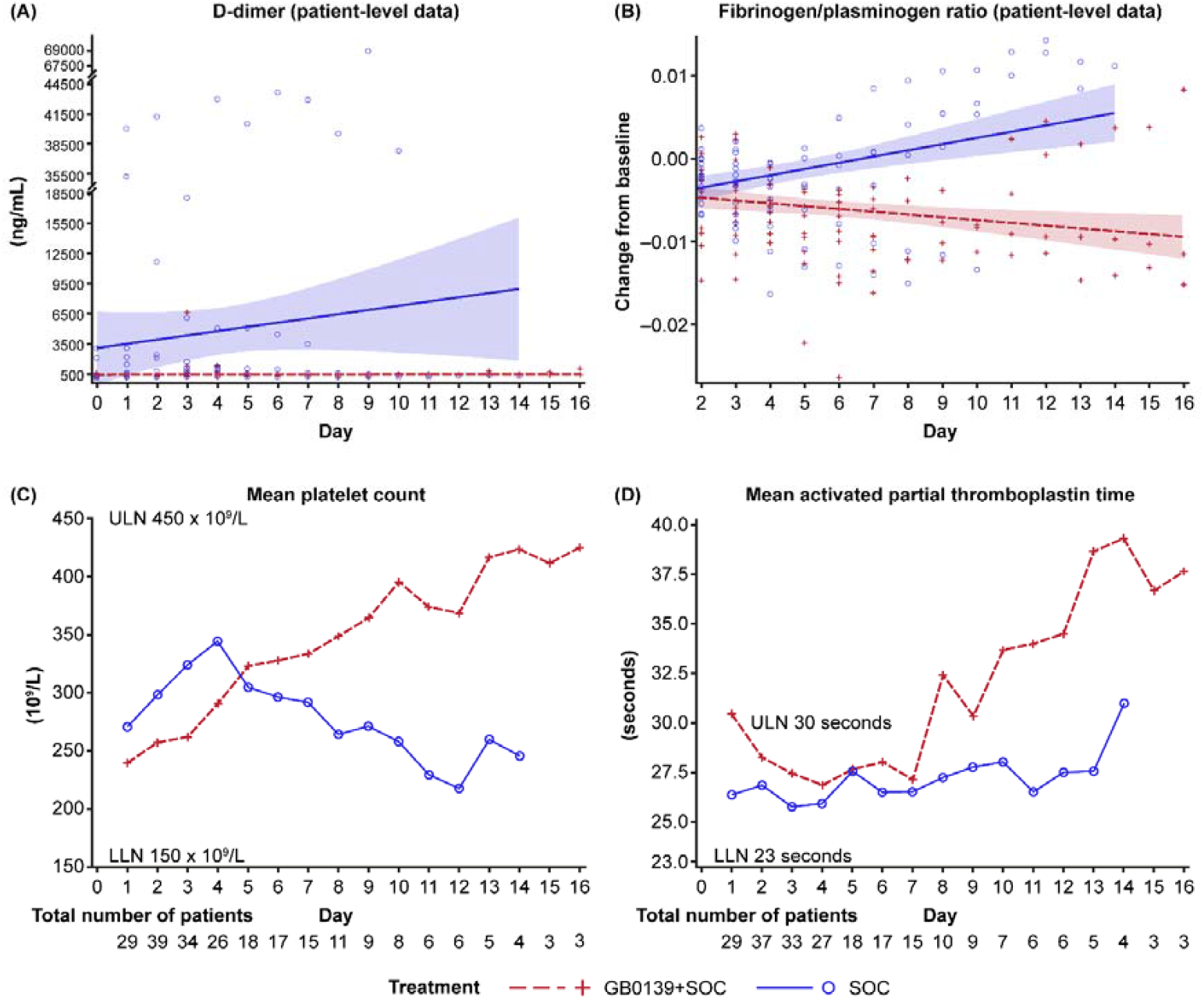
Effect of GB0139+SOC (blue) and SOC (red) on coagulopathy: individual patient-level data for (A) D-dimer levels, (B) fibrinogen/platelet ratio, (C) platelets, and (D) activated partial thromboplastin time. Parts A and B display linear regression analysis showing line of best fit with 95% CI (shaded area). CI=confidence interval. LLN=lower limit of normal. SOC=standard of care. ULN=upper limit of normal.

#### Biomarkers of organ function

Despite initial FiO_2_ being higher on admission in the GB0139+SOC arm than the SOC arm (44·05% *vs* 34·05%, respectively), the rate of FiO_2_ (% per day) decline was significantly greater with GB0139+SOC, with a posterior mean difference of -1·51 (95% HPD -2·90, -0·189) versus SOC (figure 7A). This decline was particularly evident over days 1–7 where mean FiO_2_ percentage decreased in patients treated with GB0139+SOC but increased in patients receiving SOC (figure 7B). A mild transaminitis and alterations in the levels of organ inflammation markers such as AST, GGT, and creatine kinase were also noted, in particular in the NEWS2 ≥4 patient subgroup (figure S4–C). These markers showed greater downward trends in the GB0139+SOC treatment group versus SOC, though all were within the normal range except for GGT in the SOC arm, which rose above normal levels.

**Figure 7:**
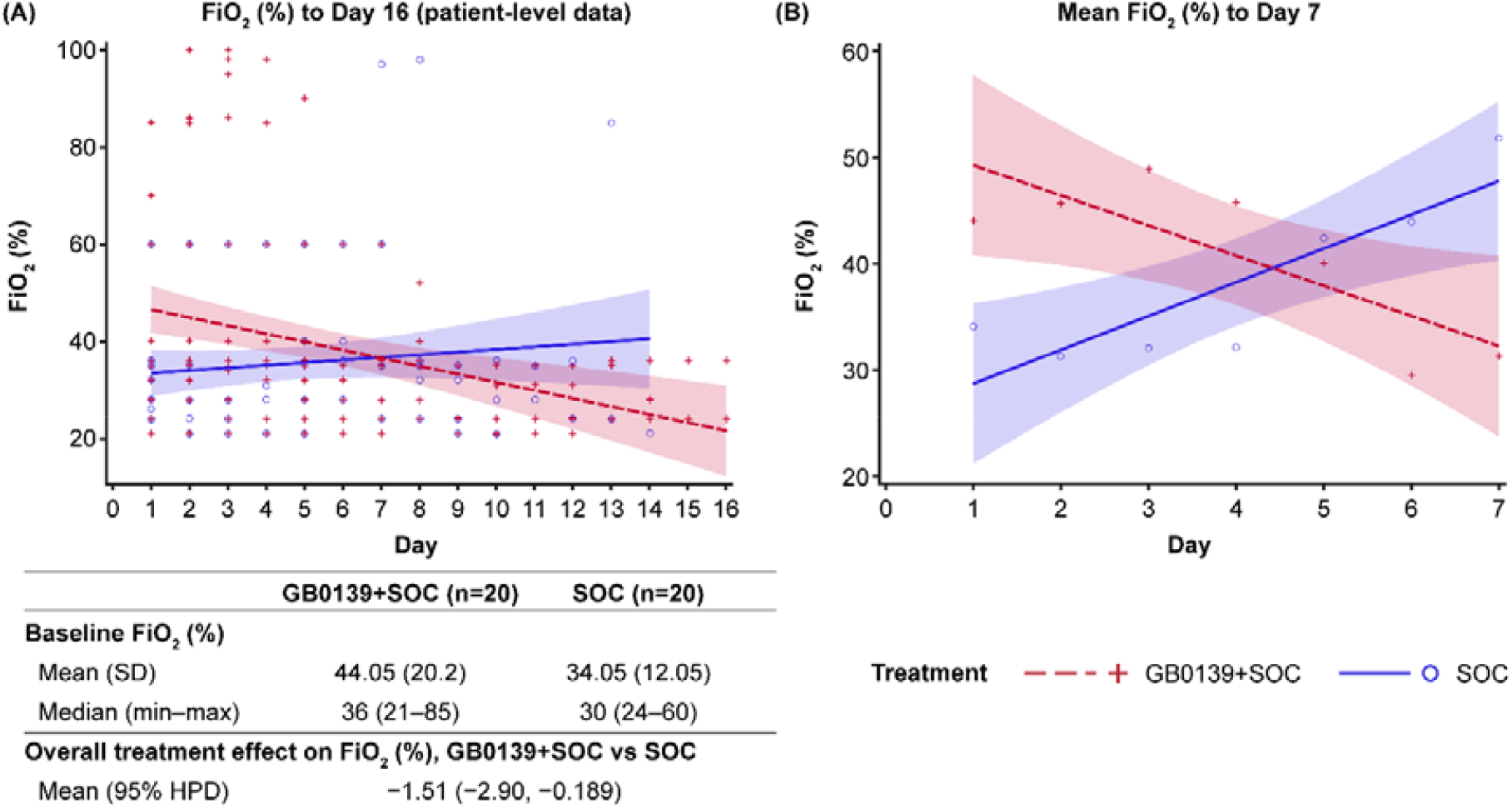
Effect of GB0139+SOC (red) and SOC (blue) on FiO_2_: (A) individual patient-level data from Days 1–16 and (B) mean FiO_2_ to Day 7. Linear regression analysis showing line of best fit with 95% CI (shaded area). CI=confidence interval. FiO_2_=fraction of inspired oxygen. HPD=highest posterior density. max=maximum. min=minimum. SD=standard deviation. SOC=standard of care.

#### Biomarkers of fibrosis

GB0139+SOC-treated patients had lowered levels (from days 4–7) of YKL-40 and exhibited a downward trend from baseline compared with SOC, where an upward trend was observed (figure 8A). In the subgroup with baseline NEWS2 ≥4, YKL-40 levels remained consistently low with GB0139+SOC, and PAI-1 was reduced over time in both treatment arms, with levels remaining lower with GB0139+SOC treatment versus SOC (figure 8A and B). Furthermore, flow analyses in this patient subgroup suggest that GB0139 may change macrophage phenotype from profibrotic transitional to antifibrotic classical and decrease transitional monocytes (figure 8C).

**Figure 8:**
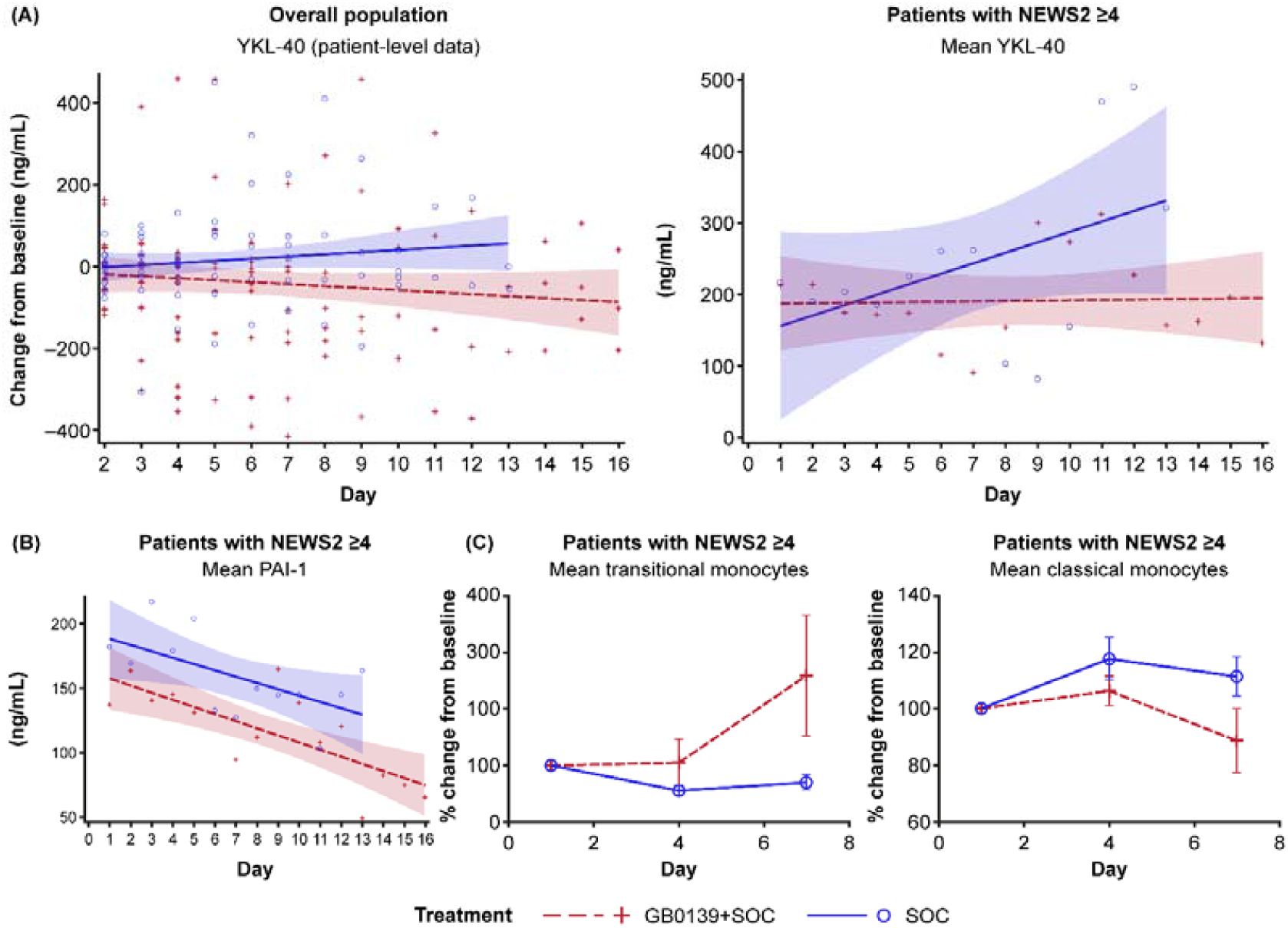
Effect of GB0139+SOC (red) and SOC (blue) on markers associated with fibrosis: (A) YKL-40, (B) PAI-1, and (C) monocyte subsets. Parts A and B display linear regression analysis showing line of best fit with 95% CI (shaded area). CI=confidence interval. NEWS2=national early warning score 2. PAI-1=plasminogen activator inhibitor-1. SOC=standard of care. YKL-40=chitinase-3-like protein 1.

### Patient mortality rates

Seven deaths occurred in the overall population of 41 randomised patients: 4 deaths in 20 patients in the GB0139+SOC arm (where patients received a median of two GB0139 doses) and 3 deaths in 21 patients in the SOC group. Six deaths occurred in the 25 patients with NEWS2 ≥4 (table S4). No deaths were attributed to GB0139.

## Discussion

This phase 2a DEFINE experimental medicine study, to our knowledge, is the first clinical trial of an inhaled galectin-3 inhibitor for the treatment of COVID-19. DEFINE met its primary endpoint with results indicating that inhaled GB0139 has an acceptable safety and tolerability profile when used in combination with SOC in hospitalised patients with COVID-19 pneumonia. AEs were limited, and no treatment-related serious AEs were reported. Despite being breathless and requiring oxygen, patients were able to inhale and achieve consistent exposure of GB0139. The 10 mg BID dose used for the first 2 days is higher than the top dose used in an ongoing study in IPF (NCT03832946), but was well tolerated. Of note, over 200 patients have been treated with GB0139 to date and none of these adverse events have been associated with GB0139 treatment. Furthermore, inhaled GB0139 caused a significant reduction of galectin-3 levels in patients with COVID-19, demonstrating target engagement, and reduced plasma levels of other key biomarkers associated with severe disease and a poor prognosis.

In the published first-in-human study of GB0139, patients with IPF were administered an inhaled dose of up to 10 mg/day for 14 days.^26^ Steady state was reached within 7 days and systemic exposure was higher in IPF patients than in healthy volunteers who received the same dose. Despite the compromised lung function with IPF, consistent delivery of GB0139 to the lung was demonstrated, with the 10 mg dose associated with an excellent PD response.^14^ Therefore, the trough concentrations for 10 mg GB0139 were used as a surrogate to gauge whether active concentrations were likely to have been reached in the lungs of patients with COVID-19 in this study. In the current study, systemic GB0139 concentrations displayed moderate interpatient variability, suggesting consistent drug delivery to the lung. We showed that inhaled GB0139 can lead to systemic exposure in COVID-19 patients comparable to that previously observed in IPF patients who subsequently had reduced levels of biomarkers associated with fibrotic disease progression.^26^ This suggests that the plasma concentrations achieved with GB0139 inhalation in patients with COVID-19 are relevant drivers of activity and a similar reduction in fibrotic biomarkers in COVID-19 could potentially decrease post-viral fibrosis.

High galectin-3 levels have been correlated with disease severity and thus is a therapeutic target in COVID-19. Patients with COVID-19 with higher serum levels of galectin-3 have greater parenchymal and vascular damage and galectin-3 levels can identify severe ARDS secondary to COVID-19 disease with good sensitivity and specificity.^32^ Furthermore, patients with high galectin-3 serum levels are not only more prone to develop severe ARDS, but are also at markedly higher risk of intensive care unit admission or death.^33^ Thus, the ability of GB0139 to reduce galectin-3 levels is encouraging.

This current study adds further weight to the therapeutic potential of galectin-3 inhibition in the treatment of patients with COVID-19. GB0139 showed a downward trend in reducing inflammation and key markers of COVID-19 severity associated with morbidity and mortality. CRP and NLR are measures of systemic inflammation, with rising levels indicative of increased severity of COVID-19 infection, and CXCL10 has been identified as a cardinal chemokine in driving COVID-19 immunopathogenesis.^34,35^ Raised CXCL10 levels are often observed in both plasma and bronchoalveolar lavage of patients who are critically ill with the disease.^36^ We have shown a trend to reduced systemic inflammation (CRP and NLR) in patients treated with GB0139+SOC compared with SOC. CXCL10 was also reduced in GB0139+SOC-treated patients versus SOC, and this was positively correlated with reduced galectin-3 up to day 4. Other key cytokines of the cytokine storm such as IL-10 and TNF also showed a greater mean downward trend from baseline following treatment with GB0139+SOC compared with SOC, and this was more evident in patients with more severe disease (baseline NEWS2 ≥4). Taken together, this suggests that GB0139, as add-on to SOC, may have the potential to further reduce the incidence of systemic inflammation and the cytokine storm in patients with COVID-19. Lymphocyte exhaustion is also a feature of both infection and progression of severe COVID-19.^37^ In the NEWS2 ≥4 subgroup, we observed that while the level of B cells and T cells may have been higher in the GB0139+SOC-treated group compared with SOC, exhaustion markers for CD4 and CD8 T cells still had a greater decrease with GB0139+SOC treatment. These observations suggest that GB0139 therapy may potentially control enhanced immunological responses to COVID-19 infection and improve recovery compared with SOC, particularly in the higher clinical risk group with NEWS2 ≥4.

A high incidence of micro/intravascular thrombosis and thromboembolic events are notable features of severe COVID-19 infection.^38^ These events can be identified by measuring coagulopathy-related biomarkers such as D-dimers, platelets, FPR, anti-thrombin III, and aPTT.^39^ In the overall population, low D-dimer levels were maintained with GB0139+SOC treatment, in contrast to SOC where a steady increase was observed. D-dimer is a fibrin degradation product and elevated concentrations (>1000 ng/mL^-1^) are strongly linked to the rate of disease complications (including severe pneumonia, hypoxia and respiratory failure, ARDS, and multiorgan failure), poor outcomes, and death.^40-44^ The trends observed in additional markers of coagulopathy (FPR, platelets and aPTT) together suggest that inhibition of galectin-3 by GB0139 administration may impact upon inflammation-induced venous thrombosis/pulmonary embolism which are major contributors to morbidity and mortality in patients with moderate-to-severe COVID-19.

Lung damage, with subsequent respiratory failure is a key feature of COVID-19,^45,46^ and FiO_2_ is an estimation of the oxygen content inhaled and thus involved in gas exchange at the alveolar level.^47^ The greater reduction in FiO_2_, observed in patients treated with GB0139+SOC compared with SOC, suggests that GB0139 improves lung function and oxygen transfer in patients with COVID-19 beyond that seen with SOC.

‘Long COVID’, particularly in relation to lung fibrosis, is an emerging long-term public health problem,^48^ and it has previously been shown that GB0139 reduces key fibrotic markers in IPF.^26^ YKL-40 is secreted by activated macrophages and neutrophils in response to IL-1 and IL-6, and associated with extracellular tissue remodelling and fibrosis and PAI-1 is a positive regulator of inflammation, clotting, and fibrosis.^49-51^ These drivers of lung fibrosis were both consistently lower following GB0139+SOC treatment versus SOC in the current analysis, particularly in patients with baseline NEWS2 ≥4. Transitional monocytes are associated with fibrosis and classical monocytes are associated with the fight against infection.^52,53^ In the NEWS2 ≥4 subgroup, we consistently observed low levels of transitional monocytes with GB0139+SOC versus SOC (<100% *vs* 100–300%) but higher overall levels of classical monocytes in the former group. Thus, GB0139 may increase the immune response to fight infection and decrease the fibrotic tendency and risk of long-term lung fibrosis in patients with moderate-to-severe COVID-19.

Mortality was comparable between the two treatment groups: four deaths in 20 patients in the GB0139+SOC arm and three deaths in 21 patients assigned to SOC. No deaths were attributed to GB0139. As previously mentioned, patients in the GB0139+SOC arm had more severe disease at baseline than those on SOC. In patients with baseline NEWS2 ≥4, there were three (21.4%) deaths amongst 14 patients who received GB0139+SOC and three (27.3%) in 11 patients receiving SOC. This amounted to a 5.9% absolute reduction in mortality rate with GB0139+SOC treatment, which was a relative reduction of 21.5%. The three patients who died in the GB0139+SOC treatment arm were treated for 2 days, 2 days, and 3 days, respectively (mean treatment time of 2.3 days). Taken together this suggests that GB0139 treatment may decrease mortality, particularly in patients with COVID-19 and high NEWS2. This, along with the improved rate of FiO_2_ decline (% per day) warrants further investigation in a larger clinical study – as this current study comprised a small population size and was not powered to detect clinically relevant differences in endpoints.

## Conclusions

DEFINE, to our knowledge, is the first trial of a galectin-3 inhibitor in COVID-19 and has demonstrated that hospitalised patients with COVID-19 can effectively inhale GB0139 and achieve plasma concentrations previously seen to induce meaningful changes in fibrotic disease-associated biomarkers. Our data indicate that GB0139 can be safely administered to patients with COVID-19 and decreases levels of galectin-3. GB0139 inhalation resulted in trends towards reduced biomarkers associated with increased morbidity and mortality in COVID-19. The consistency of the reduction in inflammatory, coagulation, and fibrosis biomarkers suggests that this is a real effect and may reduce mortality particularly in the NEWS2 ≥4 subgroup. These preliminary findings support further investigation in larger clinical trials for GB0139 as a novel therapeutic option in treating hospitalised patients with COVID-19.

## Data Availability

Ownership of the data arising from this study resides with the study team. Scientific publications and the sharing of clinical data generated as part of this trial is crucial to better understanding COVID-19 and developing new treatments. As such, the results will be submitted for publication in a peer-reviewed journal. Data will be shared, with appropriate data sharing agreements, upon request.

## Acknowledgements

This study was funded by LifeARC and funding was provided to the University of Edinburgh under the STOPCOVID award. The authors thank the participants and their families and all GB0139 study team members and investigators. The authors also thank the Clinical Research Facility staff in the Royal Infirmary of Edinburgh; the EMERGE research team; the ACCORD Facilitation, QA and Monitoring teams at University of Edinburgh/NHS Lothian (Sponsor); and Jean Antonelli for her trial management support. Thanks to Cecilia Boz, Ross Mills and Philip Emanuel for processing and analysing serum and plasma samples and to Matthew Burgess, Giulia Rinaldi, Asta Valanciute, Beth Mills, Emma Scholefield and Gareth Hardisty for sample processing and FACS collection. GB0139 is being developed by Galecto Biotech. Galecto Biotech participated in the design, study conduct, analysis, and interpretation of data, as well as the writing, review, and approval of the manuscript. Third-party medical writing assistance under the direction of the authors, was provided by Sinéad Holland, PhD, of Ashfield MedComms, an Ashfield Health company, and was funded by Galecto Biotech.

