## Supplementary materials for "GB0139, an inhaled small molecule inhibitor of galectin-3, in COVID-19 pneumonitis: a randomised, controlled, open-label, phase 2a experimental medicine trial of safety, pharmacokinetics, and potential therapeutic value"

#### SUPPLEMENTARY METHODS

##### Flow cytometry

All staining and processing were performed in a class II microbiological safety cabinet (MSCII). Centrifugation steps used capped tubes in a biosafety bucket, which were loaded and unloaded within the MSCII. Fluorescence-activated cell sorting (FACS) tubes containing fragment crystallisation (Fc) block were prepared with 5 µL of Trustain FcX (Biolegend 422301) and 5 µL of monocyte blocker (Biolegend0426102). Antibody cocktails were prepared in FACS staining buffer (phosphate buffered saline 2% flow cytometry staining buffer [Gibco]) containing Brilliant violet plus buffer (BD 566385). Five minutes after addition of 100 µL of whole blood/EDTA, 50 µL of antibody staining cocktail was added to each tube before incubation in the dark at room temperature for 20 minutes. Red blood cells were lysed using BD FACS lyse and washed twice in staining buffer before fixation (Biolegend Fixation buffer). After 20 minutes, fixed samples were moved from the MSCII to cold storage. All samples were collected on a 5 laser BD LSR Fortessa at the FACS facility at Queens Medical Research Institute within 24 hours of staining. Freshly prepared 8 peak calibration beaks were ran daily prior to sample collection.

##### Gating strategies

After exclusion of debris and selection of lymphocytes on FSC (forward scatter) versus SSC (side scatter) doublets were excluded and CD3+ T cells selected. CD4+ and CD8+ T cell subsets were selected and subdivided into naïve versus memory subsets according to expression of CD45RA and CCR7 as shown. Expression of T-cell receptor γδ on double negative cells expressing CD3 but neither CD4 nor CD8 was used to identify gamma-delta-T cells. Treg were gated within the CD4+ population as CD25+CD127-low cells.
After exclusion of debris and doublets CD45+ cells were selected and subdivided into lymphocytes, monocytes and granulocytes according to CD45 expression level and side scatter properties. Within the lymphocyte gate B cells were selected as CD19+ and antibody secreting cells identified as CD27+CD38+. Lineage negative monocytes (CD3/CD19/CD66b/CD56−) were subdivided to classical, transitional and non-classical subsets according to expression of CD14 and CD16. Granulocytes were subdivided into neutrophils and eosinophils on the basis of CD16 expression.
After exclusion of debris and doublets SSC-H high, lineage negative (CD3/CD19/CD56/Siglec-8) events were gated and CD15+CD66b+ cells selected as neutrophils. Immature neutrophils were gated as CD10 low.

#### SUPPLEMENTARY TABLES

##### Table S1: List of antibodies

| Target | Fluorophore | Clone | Isotype | Category number |
|---|---|---|---|---|
| CD3 | AF700 | UCHT1 | ms IgG1 | 300424 |
| CD4 | BV711 | OKT4 | ms IgG2b | 317440 |
| CD8 | BV421 | RPA-T8 | ms IgG1 | 301036 |
| CD57 | BV605 | QA17A04 | ms igG1 | 393304 |
| CD45RA | FITC | HI100 | ms IgG2b | 304148 |
| CCR7 | PE-Dazzle | GO43H7 | ms IgG2a | 353236 |
| CXCR3 | PE | GO25H7 | ms IgG1 | 353706 |
| CCR6 | PerCP-Cy5.5 | G034E3 | ms IgG2b | 353406 |
| CD103 | APC | BerACT8 | ms IgG1 | 350216 |
| CD56 | APC-cy7 | HCD56 | ms IgG1 | 318332 |
| CD45RO | PerCP | UCHL1 | ms IgG2a | 304252 |
| TCR gamma delta | BV510 | B1 | Ms IgG1 | 331220 |
| CD38 | FITC | HIT2 | ms IgG1 | 303504 |
| HLA-DR | APC | L243 | ms IgG2a | 307610 |
| CD39 | BV605 | A1 | ms IgG1 | 328236 |
| PD-1 | PE | EH12.2H7 | ms IgG1 | 329905 |
| CD25 | BV650 | V T0-72 | ms IgG2b | 302634 |
| CD127 | PerCP-cy5.5 | A019D5 | ms IgG1 | 351321 |
| Tim3 | PeDazzle | F38-2E2 | ms IgG1 | 354034 |
| CD45 | PerCP | 2D1 | ms IgG1 | 368506 |
| CD3 | AF700 | SK7 | ms IgG1 | 344822 |
| CD19 | PE-dazzle | HIB19 | ms IgG1 | 302252 |
| CD56 | APC-cy7 | HCD56 | ms IgG1 | 318332 |
| CD14 | BV711 | 63D3 | msIgG1 | 367140 |
| CD16 | BV421 | VNK80 | msIgG1 | 302038 |
| HLA-DR | APC | L243 | ms IgG2a | 307610 |
| CD66b | PE | QA17A51 | ms IgG1 | 392904 |
| CD27 | PerCP-cy5.5 | M-T271 | ms IgG1 | 356408 |
| CD38 | FITC | HIT2 | ms IgG1 | 303504 |
| CD274 | BV650 | 29E.2A3 | ms IgG2b | 329740 |
| HLA-DR | BV650 | L243 | ms IgG2a | 307650 |
| CCR2 | PerCPcy5.5 | K03632 | Ms IgG2a | 357204 |
| CX3CR1 | PE-dazzle | 2A9-1 | rat igG2b | 341624 |
| CCR5 | APC-cy7 | J418F1 | rat igG2b | 359110 |
| CD3 | APC | UCHT1 | ms IgG1 | 300412 |
| CD19 | APC | HIB19 | ms IgG1 | 302212 |
| CD56 | APC | HCD56 | ms IgG1 | 318310 |
| Siglec8 | APC | 7C9 | msIgG1 | 347106 |
| CD66b | PE | 6/40c | ms IgG1 | 392904 |
| CD15 | APC-Cy7 | W6D3 | msIgG1 | 323048 |
| CD11b | PerCPcy5.5 | LM2 | ms IgG1 | 393106 |
| CD62L | PE-Dazzle | DREG-56 | ms IgG1 | 304842 |
| CD64 | BV711 | 10.1 | ms IgG1 | 305042 |
| CD16 | BV421 | 3g8 | msIgG1 | 302038 |
| CD10 | BV605 | HI10a | msIgG1 | 312222 |
| CD54 (ICAM-1) | AF700 | HA58 | ms IgG1 | 353126 |
| CD181 (CXCR1) | AF488 | 8F1 CXCR1 | ms IgG2a | 320616 |

All antibodies were supplied by Biolegend.

**Table S2: Continuous outcome variable results: haematological and biochemical safety laboratory outcomes and vital signs**

| Variable | Mean | 95% HPD interval | |
|---|---|---|---|
| | | Lower limit | Upper limit |
| Haemoglobin (g/L) | −0·495 | −7·21 | 5·963 |
| Haematocrit (ratio) | −0·206 | −5·04 | 4·996 |
| Red cell count (109/L) | −0·128 | −4·87 | 4·641 |
| Mean cell volume (fl) | −0·136 | −5·73 | 5·546 |
| Mean cell Hb (pg) | −0·344 | −5·36 | 4·608 |
| Mean cell Hb concentration (g/L) | 0·050 | −6·94 | 7·375 |
| White cell count ($10^9$/L) | −0·489 | −6·04 | 5·255 |
| Neutrophil count ($10^9$/L) | 0·031 | −5·80 | 6·003 |
| Lymphocytes ($10^9$/L) | −0·030 | −5·64 | 5·422 |
| Monocyte count ($10^9$/L) | 0·112 | −5·93 | 5·751 |
| Basophil count ($10^9$/L) | −0·640 | −5·88 | 4·626 |
| Eosinophil count ($10^9$/L) | 0·691 | −10·8 | 12·99 |
| Platelet count ($10^9$/L) | 17·76 | −25·5 | 59·05 |
| Random glucose (micromole/L) | −0·536 | −6·45 | 4·911 |
| Urea (mmol/L) | 0·533 | −5·08 | 6·125 |
| Sodium (mmol/L) | −0·203 | −5·25 | 5·327 |
| Potassium (mmol/L) | 0·144 | −4·96 | 5·791 |
| Chloride (mmol/L) | −1·05 | −7·05 | 4·631 |
| Magnesium (mmol/L) | −0·090 | −5·95 | 6·038 |
| Bicarbonate (mmol/L) | 0·856 | −4·65 | 6·472 |
| Creatinine (micromole/L) | −1·77 | −8·39 | 4·721 |
| Total protein (g/L) | −0·896 | −6·93 | 5·036 |
| Albumin (g/L) | −1·05 | −6·37 | 4·081 |
| AST (U/L) | −10·3 | −28·4 | 7·814 |
| Total bilirubin (micromole/L) | 0·741 | −4·83 | 6·798 |
| GGT (U/L) | −15·2 | −44·2 | 15·82 |
| ALT (U/L) | −2·97 | −26·2 | 20·77 |
| Alkaline phosphatase (U/L) | −1·37 | −9·75 | 6·990 |
| LDH (U/L) | −18·8 | −62·7 | 25·53 |
| Ferritin (micromole/L) | 63·66 | −96·9 | 220·1 |
| CRP (mg/L) | 0·168 | −27·1 | 27·82 |
| Troponin (ng/L) | −0·469 | −10·5 | 10·06 |
| Triglycerides (mmol/L) | −0·696 | −6·69 | 4·959 |
| D-dimer (ng/ml) | 5·609 | −208 | 209·1 |
| Creatinine kinase (U/L) | −23·1 | −76·8 | 25·12 |
| INR (ratio) | −0·444 | −5·43 | 4·268 |
| Prothrombin time (seconds) | −0·484 | −5·42 | 4·740 |
| Fibrinogen (g/L) | −0·071 | −6·14 | 5·822 |
| aPTT (seconds) | 0·226 | −6·06 | 6·674 |

| | | | |
|---|---|---|---|
| Protein C (IU/ml) | −0·134 | −5·78 | 5·671 |
| Antithrombin (IU/ml) | −0·131 | −6·16 | 5·718 |
| $O_2$ saturation (%) | 0·616 | −3·49 | 4·848 |
| Respiratory rate (bpm) | 0·774 | −5·06 | 6·715 |
| Systolic Blood Pressure (mm Hg) | −3·62 | −13·6 | 6·819 |
| Diastolic Blood Pressure (mm Hg) | −0·773 | −7·81 | 6·425 |
| Temperature (degrees C) | 0·196 | −4·78 | 5·035 |
| Pulse (bpm) | 5·242 | −4·52 | 14·52 |
| ECG rate | 2·143 | −8·11 | 12·86 |
| ECG QTc | −4·37 | −22·5 | 13·09 |

All of the 95% HPD credible intervals include zero, and therefore none are statistically significant. Some posterior means may be of a sufficiently high magnitude to be clinically relevant. ALT=alanine aminotransferase. aPTT=activated partial thromboplastin time. AST=aspartate aminotransferase. CRP=c-reactive protein. ECG=electrocardiogram. GGT=gamma-glutamyltransferase. Hb=haemoglobin. HPD=highest posterior density. INR=international normalised ratio. LDH=lactate dehydrogenase. QTc=corrected QT interval.

**Table S3: Trough (pre-dose) plasma GB0139 concentrations (ng/mL) in IPF patients (unpublished data on file from ClinicalTrials.gov identifier: NCT02257177)**

| | Day 2 | Day 3 | Day 7 | Day 14 | Day 15 |
|---|---|---|---|---|---|
| n | 5 | 5 | 5 | 5 | 5 |
| Mean (SD) | 12·93 (7·88) | 16·02 (9·83) | 14·88 (7·63) | 16·38 (8·56) | 14·66 (8·64) |
| CV, % | 61 | 61 | 51 | 52 | 59 |
| Geometric mean | 10·21 | 13·25 | 12·77 | 13·53 | 12·17 |
| Median (min–max) | 14·99 (2·55–22·49) | 15·11 (4·24–30·15) | 15·19 (4·16–23·81) | 15·33 (3·42–24·74) | 15·18 (3·95–26·14) |

CV=coefficient of variation. IPF=idiopathic pulmonary fibrosis. max=maximum. min=minimum. SD=standard deviation.

**Table S4: Mortality in patients with COVID-19 with NEWS2 ≥4 in DEFINE**

| Treatment | No. of patients | Deaths (%) | Narrative |
|---|---|---|---|
| GB0139+SOC | 14 | 3 (21·4) | Male; aged 71–75 years; BMI of 29·3; comorbid cardiovascular and lung disease; GB0139 received over 2 days; died within 1 week of hospital admission<br><br>Male; aged 86–90 years; BMI of 33·3; comorbid cardiovascular disease; initially treated with SOC, heparin, dexamethasone, antibiotics, GB0139 received over 2 days; the patient was extremely unwell since admission with very high oxygen requirements; died within 10 days of hospital admission<br><br>Male; aged 61–65 years; BMI of 29; comorbidities present including cardiovascular and metabolic disease; GB0139 received over 3 days (NEWS2 decreased from 5 to 3); died within 3 weeks of hospital admission |
| SOC | 11 | 3 (27·3) | Male; aged 81–85 years; BMI of 32·7; comorbid cardiovascular, lung, and metabolic disease; died within 10 days of hospital admission<br><br>Female; aged 61–65 years; BMI of 38·6; comorbid cardiovascular and lung disease; died within 3 weeks of hospital admission<br><br>Female, aged 81–85 years; BMI of 28·8; comorbid cardiovascular and inflammatory disease; short admission |

BMI=body mass index. NEWS2=National Early Warning Score 2. SOC=standard of care.

### SUPPLEMENTARY FIGURES

#### Figure S1: Gating strategies

Gating strategies for T cell subsets

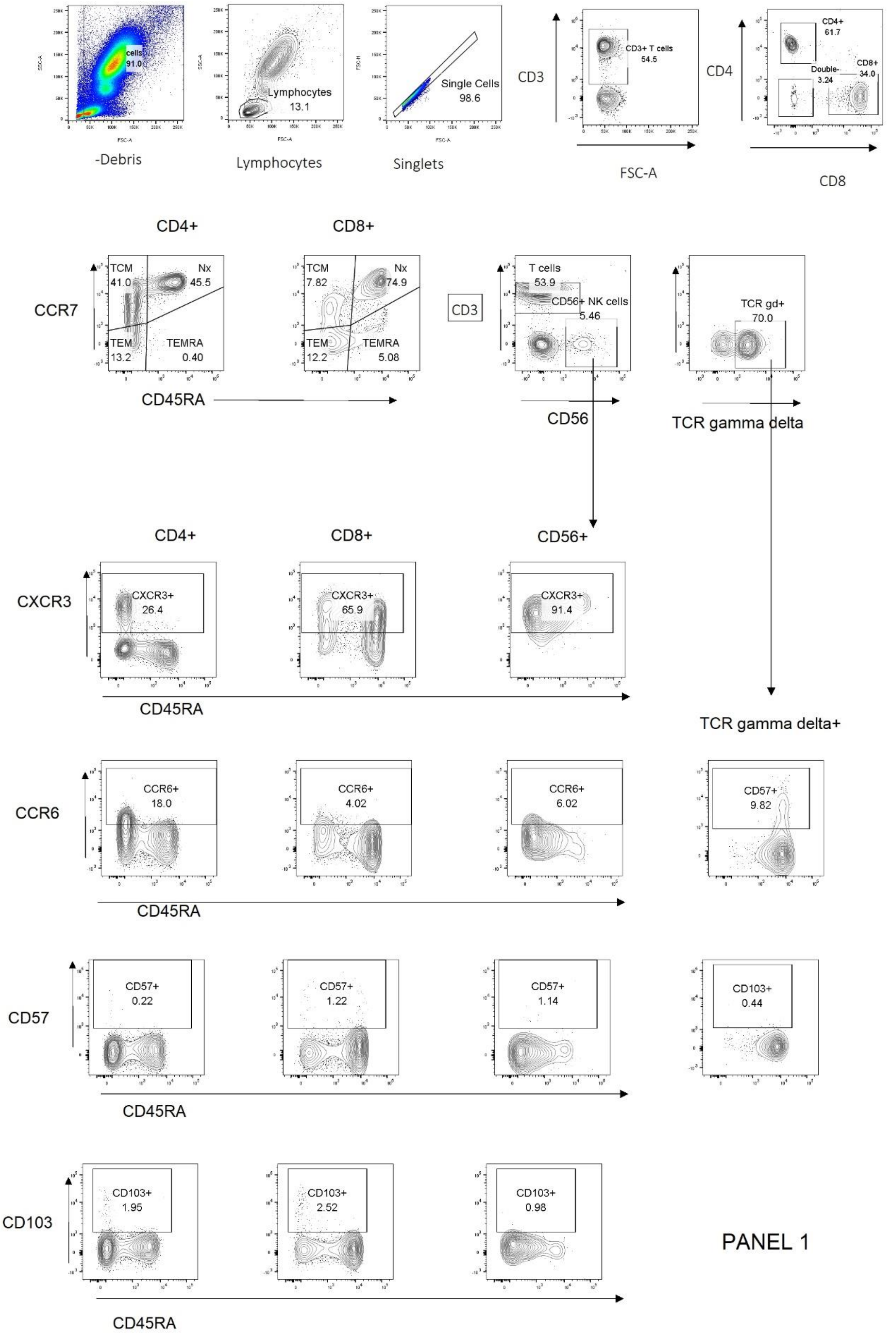

Gating strategy for T cell subsets

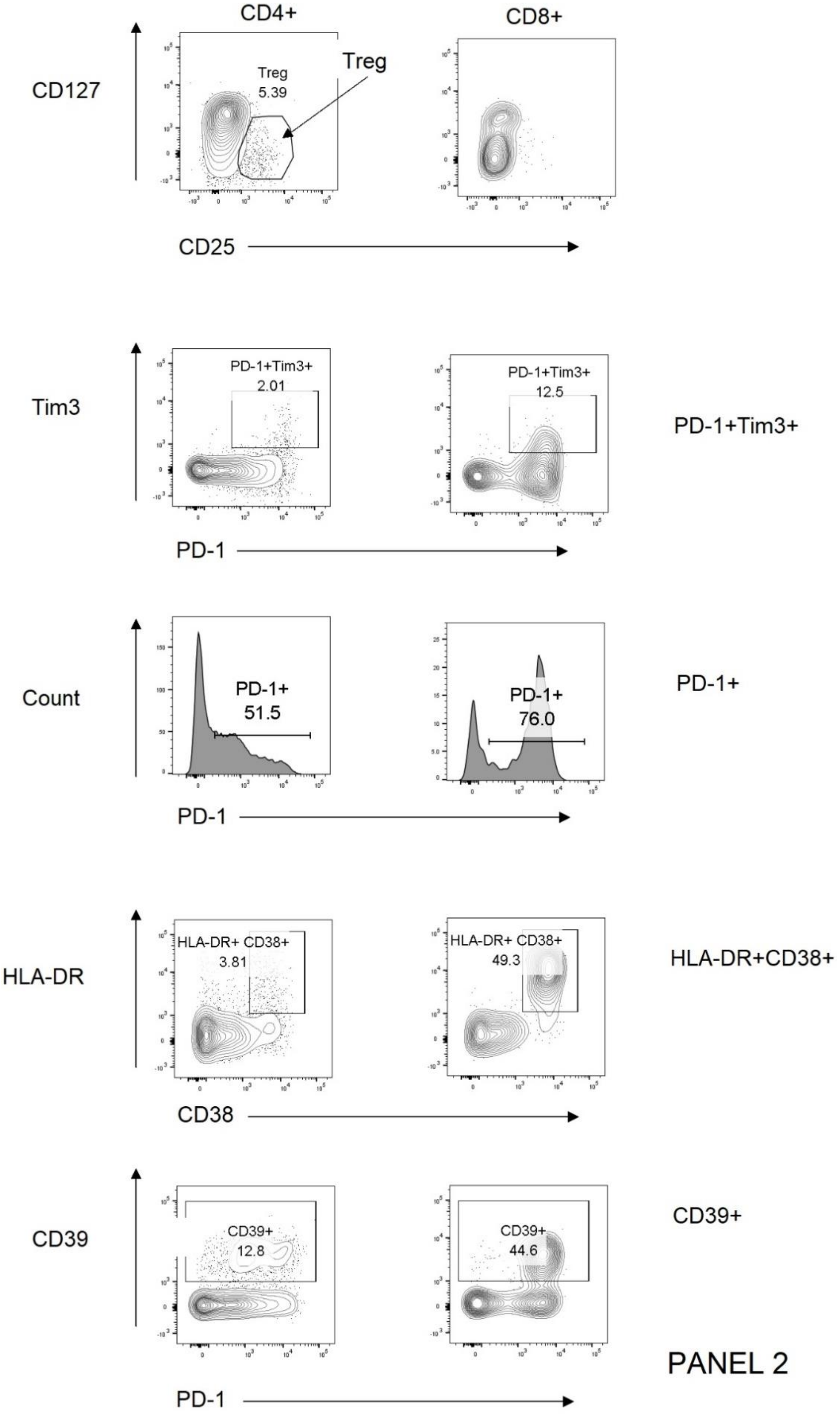

Gating strategy for B cells and monocytes

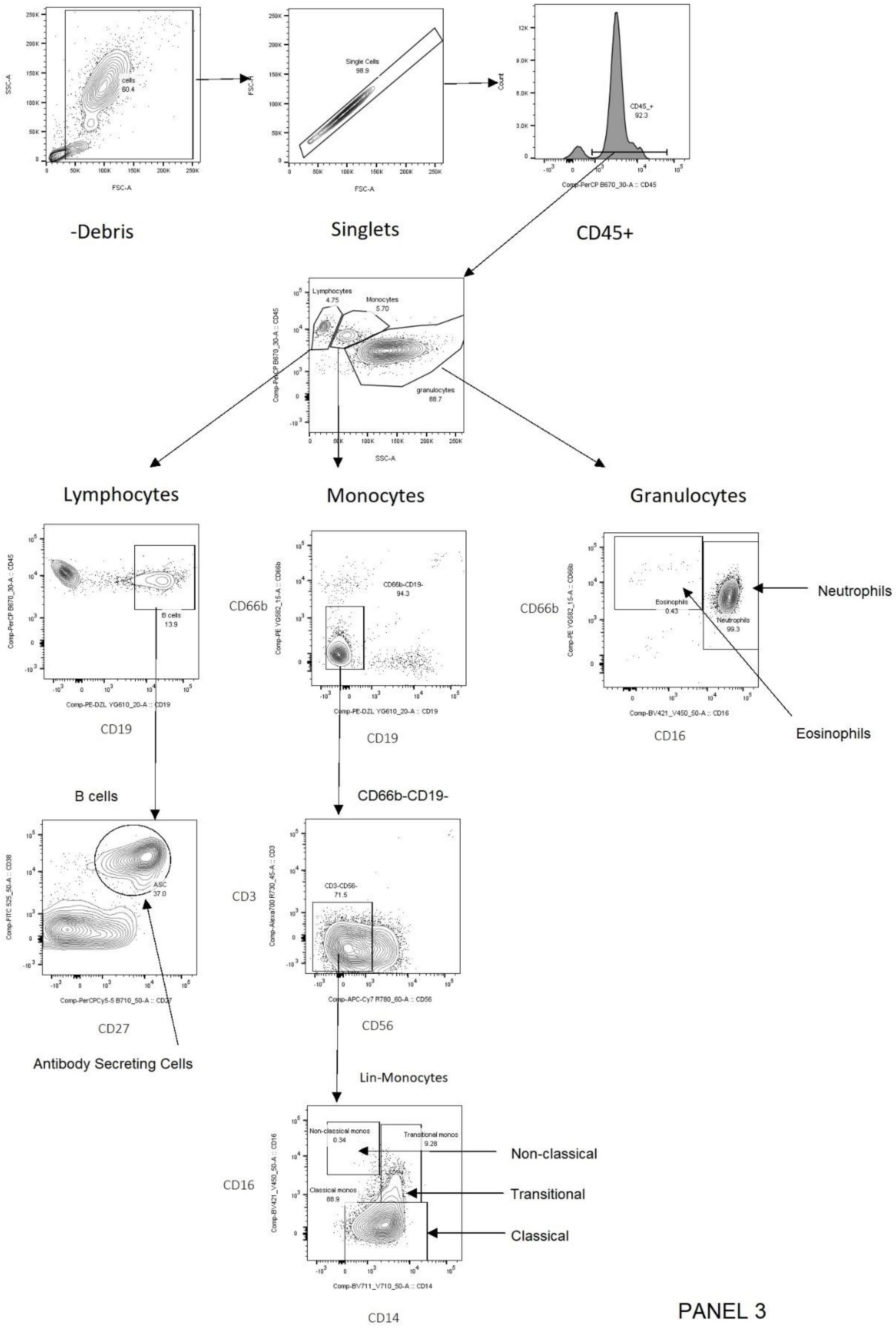

Gating strategy for neutrophils

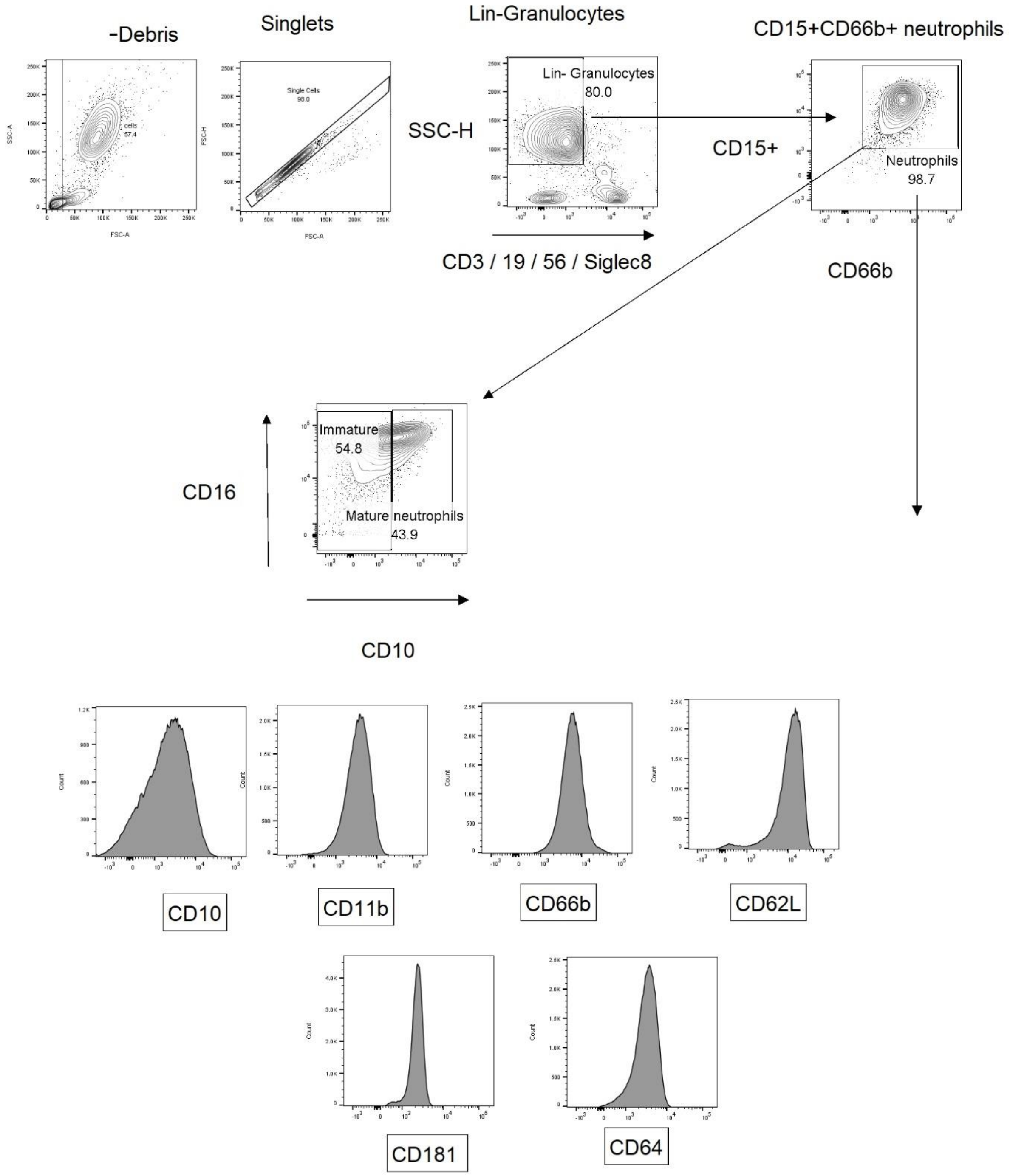

**Figure S2: GB0139 plasma concentrations in patients with COVID-19 and patients with IPF**

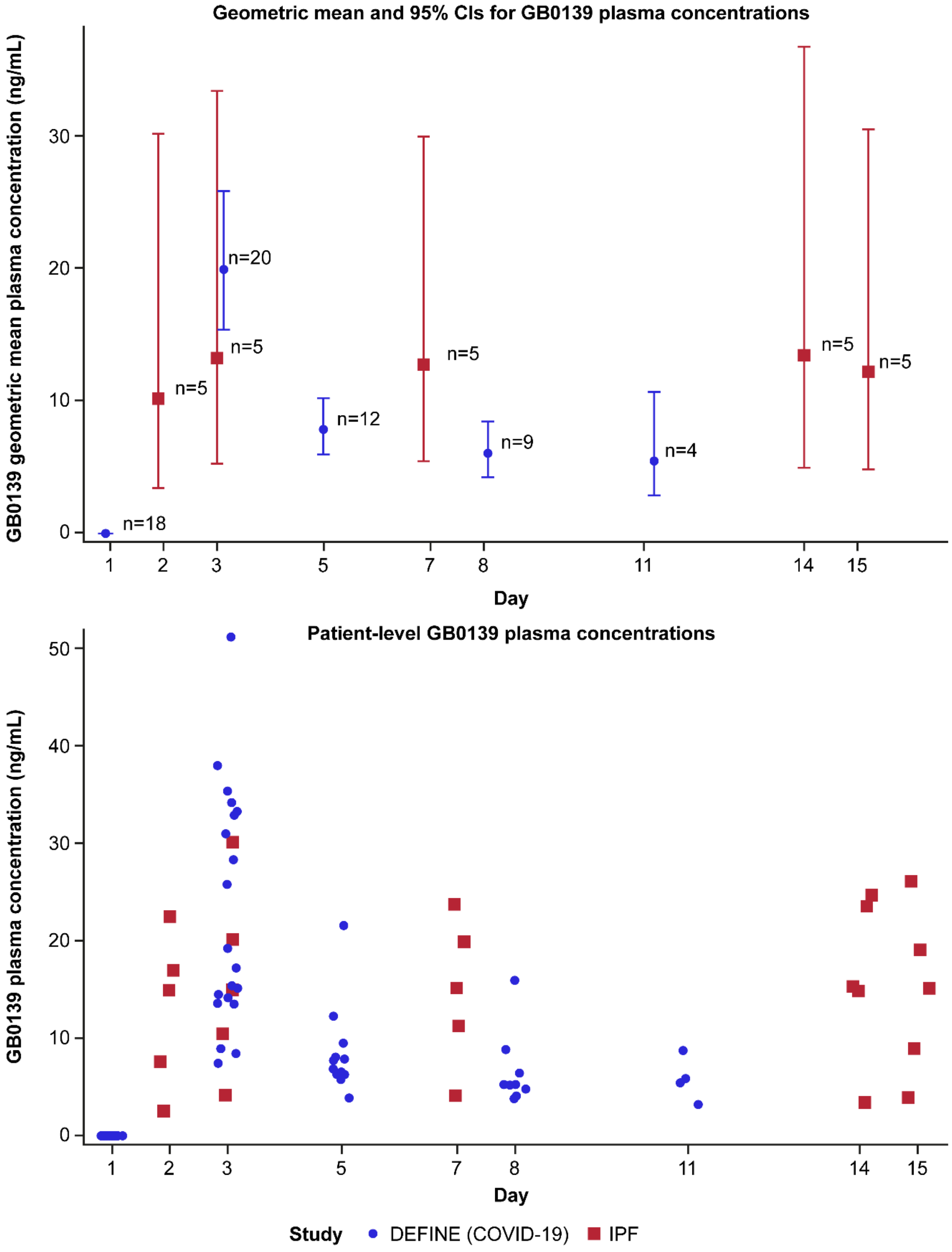


CI=confidence interval. IPF=idiopathic pulmonary fibrosis.

**Figure S3: Effect of GB0139+SOC (red) and SOC (blue) on markers of inflammation in the overall population: (A) IL-10, (B) IL-6, and (C) tumour necrosis factor**

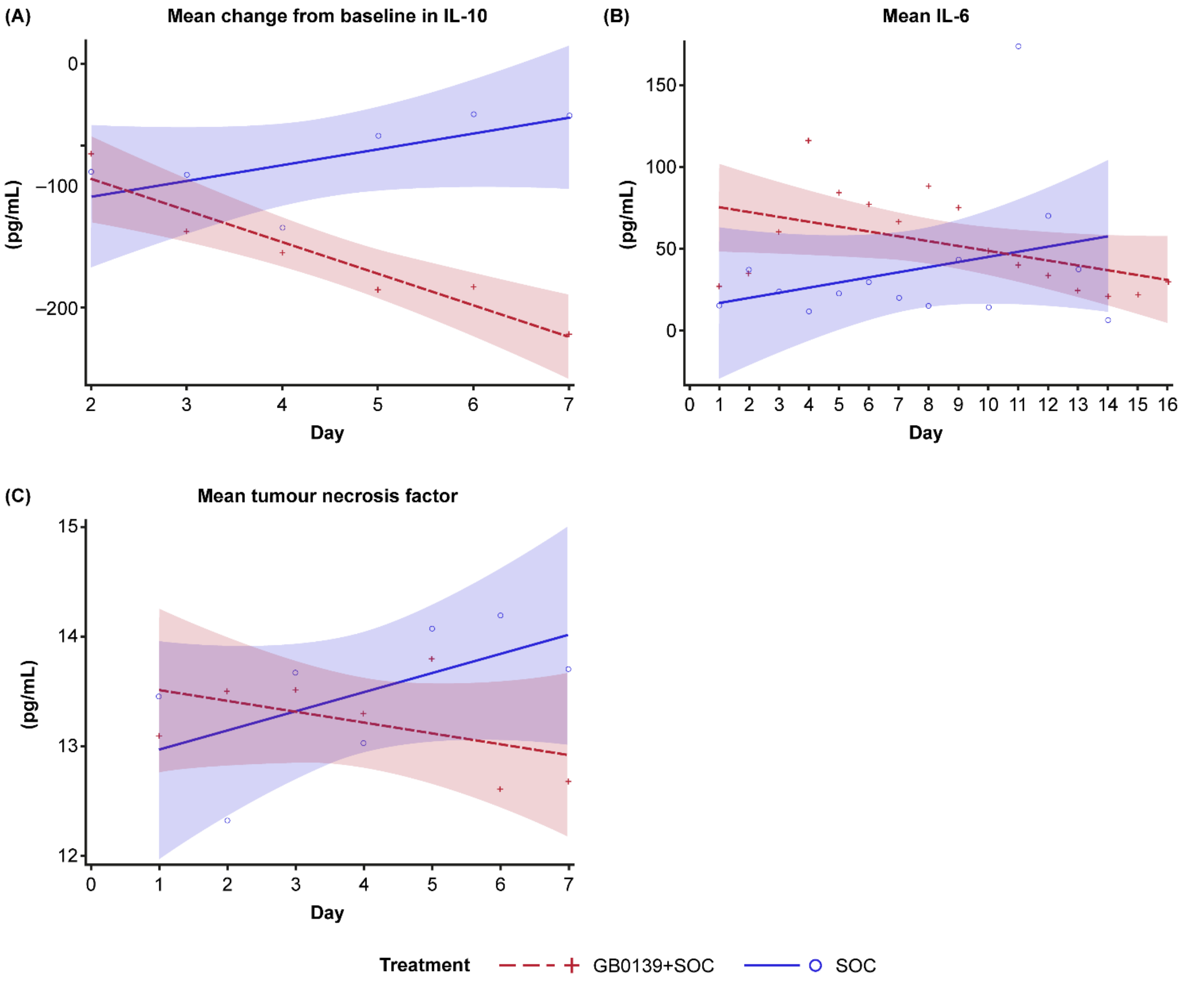


Linear regression analysis showing line of best fit with 95% CI (shaded area). CI=confidence interval. IL=interleukin. SOC=standard of care.

**Figure S4: Treatment with GB0139+SOC indicates favourable trends, versus SOC alone, in organ function preservation in patients with NEWS2 ≥4: (A) liver function (aspartate aminotransferase, gamma-glutamyl transpeptidase) and (B) general tissue damage (creatine kinase)**

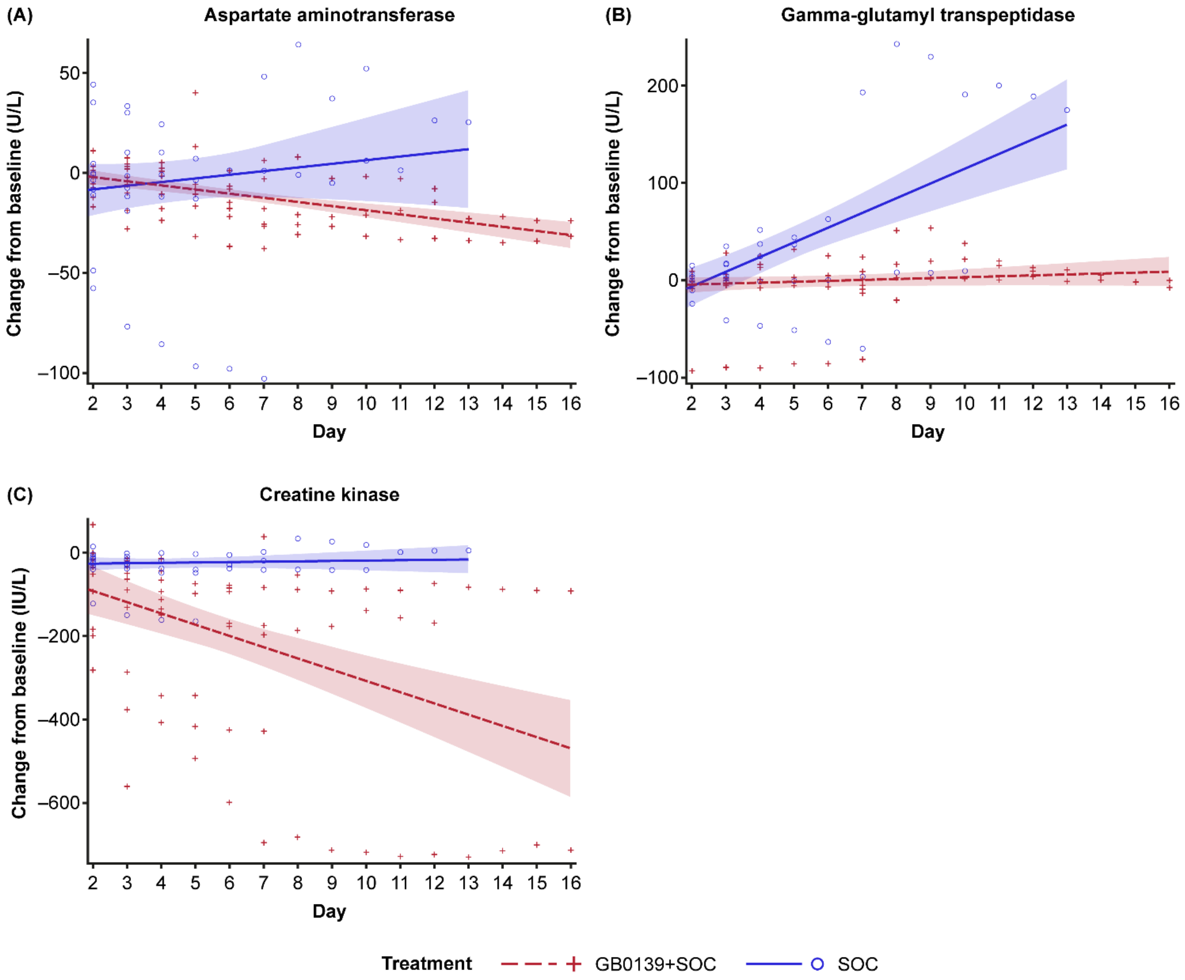


Linear regression analysis showing line of best fit with 95% CI (shaded area). CI=confidence interval. NEWS2=national early warning score 2. SOC=standard of care.
